# The potential clinical impact and cost-effectiveness of the updated COVID-19 mRNA Autumn 2024 vaccines in the United Kingdom

**DOI:** 10.1101/2024.08.19.24312243

**Authors:** Michele A. Kohli, Michael Maschio, Amy Lee, Keya Joshi, Stuart Carroll, Orsolya Balogh, Nicholas van de Velde, Ekkehard Beck

## Abstract

**Aims:** To estimate the potential clinical impact and cost-effectiveness of a United Kingdom (UK) Autumn 2024 vaccination campaign with an updated Moderna COVID-19 vaccine in adults ≥65 years and eligible persons 6 months to 64 years of age over a 1-year time horizon (September 2024-August 2025).

**Materials and methods:** A compartmental Susceptible-Exposed-Infected-Recovered model was adapted to reflect COVID-19 cases in the UK. Numbers of symptomatic infections, COVID-19– related hospitalizations and deaths, costs, and quality-adjusted life-years (QALYs) were predicted using a decision tree. The incremental cost-effectiveness ratio (ICER) of an updated Moderna mRNA vaccine (Moderna Autumn 2024 Campaign) was compared to no Autumn 2024 vaccine and to an updated Pfizer-BioNTech mRNA Autumn 2024 vaccine, from a healthcare perspective.

**Results:** The Moderna Autumn 2024 Vaccination Campaign is predicted to decrease the expected 8.3 million symptomatic infections with no vaccination by 19% to 6.7 million. Hospitalizations, long COVID cases, and deaths are expected to decline by 27,000 (−38%), 59,000 (−19%), and 6000 (−43%), respectively. The Moderna Autumn 2024 Campaign will increase QALYs by 78,000 and costs by £665 million, yielding an ICER of £8500/QALY gained. Sensitivity analyses suggest that vaccine effectiveness (VE) and waning, symptomatic infection incidence, hospitalization rates, and mortality rates drive cost-effectiveness. Vaccination remains cost-effective when lowering the target population to ≥50 years. Use of the Moderna vaccine is expected to prevent 8000 more hospitalizations and 1700 more deaths than the updated Pfizer-BioNTech vaccine.

**Conclusions:** Vaccination of the eligible population would contribute to significant reductions in hospitalizations, deaths, and long COVID in the UK in the 2024-2025 season. Expanding the target population continues to be cost-effective. Use of the Moderna Autumn 2024 Campaign is predicted to reduce SARS-CoV-2 infections and associated outcomes in a cost-effective manner and will contribute to a more resilient healthcare system in the UK.

## Introduction

COVID-19 continues to pose a substantial healthcare and societal burden in the United Kingdom (UK). According to the UK Health Security Agency (UKHSA), there were almost 120,000 COVID-19 patients admitted to hospital, and 9,700 mortalities with COVID-19 listed on the death certificate between July 2023-June 2024.[1,2] COVID-19 related hospital admission has been associated with higher long-term risks of death and adverse outcomes in several organ systems, especially in older adults.[3] In the whole population, the virulence and transmissibility of emerging variants continue to pose significant risk through pressure on the healthcare system and productivity loss, especially for those who are not routinely vaccinated.[4–6] One of the stated goals of the Joint Committee on Vaccination and Immunisation (JCVI) is to reduce the burden of COVID-19 on the National Health Service (NHS), especially during the winter months when other respiratory infections are expected to achieve peak incidence.[7] Therefore, another facet of the burden of COVID-19 hospitalizations, are the bed days taken by COVID-19 patients that would have been allocated for other required hospitalizations. In addition to the acute SARS-CoV-2 infection, there is the risk of post-COVID syndromes (including long COVID)[8] which can have prolonged and debilitating effects on individuals and place a substantial burden on the healthcare system. Thus, the health and economic impact of COVID-19, including costs, health-related quality-of-life, and healthcare system capacity associated with both acute infection and sequelae following acute infection, as well as ways to reduce this burden, such as the impact of vaccination on the risk, remain areas of considerable interest.[9,10]

The UK conducted multiple spring and autumn booster campaigns between the years 2022 and 2024, targeting those considered at most risk of severe outcomes following infection, based on eligibility criteria recommended by the JCVI.[11] The 2022 autumn booster campaign targeted all adults aged 50 years and over, as well as those aged 5-49 at risk for more severe outcomes, their carers, and health and social care workers.[12] This population was narrowed for the 2023 autumn booster campaign, with vaccination being recommended only for all adults aged 65 years and older and for those at high risk of COVID-19 (aged 6 months to 64 years), their carers, and health and social care workers.[7] Target populations for Spring 2023 and Spring 2024 booster campaigns were even narrower, with only adults aged 75 years, residents of older adult care homes, and immunosuppresed recommended for the additional boosters.[13,14] The recent recommendations from the JCVI for the autumn 2024 booster recommend the vaccination of adults aged 65 years and older, residents of care homes and persons aged 6 month to 64 years at risk for more severe outcomes.[15] The NHS is also offering vaccines to frontline health and social care workers and staff in care homes for older adults in Autumn 2024.[16]

The monovalent XBB variant versions of the Moderna and Pfizer-BioNTech mRNA COVID-19 vaccines were used for the Autumn 2023 and Spring 2024 vaccination campaigns.[11] Both of these vaccines, with versions that contain updated variants, are expected to be used in the Autumn 2024 campaign.[15]

Although the Moderna and Pfizer-BioNTech COVID-19 vaccines are both based on mRNA technology, there are differences in their formulations, including dose and lipid nanoparticle mRNA delivery system.[17–20] Observational studies have found Moderna formulations used in previous vaccination campaigns to result in higher vaccine effectiveness against infections and hospitalizations than the Pfizer-BioNTech formulations.[21] A meta-analysis by Kavikondala et al, (2024) which included 24 observational studies, found a significantly lower risk of infections (including symptomatic and severe infections, risk ratio [RR] for infection 0.72 [95% CI 0.64-0.80]) and hospitalization 0.65 (95% CI 0.53-0.79)) associated with the Moderna vaccine compared to the Pfizer-BioNTech vaccine in older adults aged ≥50 years.[22] Therefore, choice and availability of COVID-19 mRNA vaccines, particularly in the upcoming tendering environment in the UK in 2025, becomes a relevant question.

Despite the availability of effective vaccines, SARS-CoV-2 continues to cause infections year-round with the protective effect of the vaccine varying based on circulating variants and time since vaccination. In light of this continued risk of infection to the population, it is important to devise vaccination strategies that ensure direct protection of the most vulnerable, while recognizing the remaining uncertainties and changes in COVID-19 epidemiology over time, such as risk of high transmission and rapid surge in infection rates. Future vaccine strategies must, therefore, be guided by the most current evidence, considering epidemiology and both clinical and economic aspects of additional vaccine doses, and must strategically focus on protecting those most at risk of severe illness. Therefore, the objectives of this study were multi-fold. Firstly, to determine, with the use of mathematical modeling, the potential cost-effectiveness of continuing a 2024 Autumn Vaccination Campaign using a Moderna variant-updated vaccine (Moderna Autumn 2024 Campaign) compared to no vaccination campaigns (No Autumn 2024 Campaign) over a one-year time horizon (September 1, 2024-August 31, 2025), assuming a similar target population as in the 2023 Autumn Campaign. This target population is very similar to the recently released NHS recommendations for the 2024 Autumn Campaign, which included adults ≥65 years plus individuals 6 months to 64 years who are at clinical risk of severe outcomes as well as boosters being offered to all residents and staff of care homes for older adults, and frontline health and social care workers.

Secondly, the cost-effectiveness of expanding the target population to include those 50 years and older was examined. The third objective was to compare the clinical and economic impact of using a variant-updated Moderna vaccine (Moderna Autumn 2024 Vaccine) compared to a variant-updated Pfizer-BioNTech vaccine (Pfizer-BioNTech Autumn 2024 Vaccine).

## Methods

### Overview

A previously developed Susceptible-Exposed-Infected-Recovered (SEIR) dynamic transmission model and infection consequence decision tree model, described in detail by Kohli et al. (2023) for the United States,[23,24] was adapted to the UK for this analysis. The SEIR model was used to predict the total number of infections (i.e., asymptomatic and symptomatic) and residual protection due to prior vaccinations during the analytic time horizon. These values were entered into the infection consequences decision tree to estimate the numbers of symptomatic infections, COVID-19–related hospitalizations and deaths, and associated costs and quality-adjusted life-years (QALYs) losses for each vaccination strategy. The number of adverse events (AEs) and their associated costs and QALYs lost were also estimated for each vaccine strategy. The UK-specific adaptations and data are described below and in the Technical Appendix. Further information on the base model structure have been previously presented.[23,24]

### SEIR dynamic transmission model

The adapted SEIR model, presented in Figure 1, consists of multiple strata representing the different vaccine campaigns that have been previously carried out in the UK. The black arrows represent the force (incidence) of infection which moves individuals from Susceptible (S) to Exposed (E) states; while in the E states, infections are both asymptomatic and nontransmissible. The force of infection is a function of the number of susceptible people in the UK population as well as the rate of effective contacts between individuals in the S and Infected (I) states. The average daily vaccine effectiveness (VE) reduces the probability of transmission in each vaccine strata. After an average of 3 days,[25] individuals move from the E state to the I state (blue arrows), which represents a transmissible infection that may be clinically symptomatic or asymptomatic. After an average of 7 days,[25,26] individuals then move to the Recovered (R) states where they have natural immunity due to the recent infection. Individuals move from the R to S states as natural immunity wanes. Once individuals have received their primary series, they move into the “Vaccinated” stratum. They progress through the booster strata over time (e.g. Booster 1, Spring 2022 booster, Autumn 2022 booster, etc.) based on the timing of the booster as additional doses are received. Individuals in any S or R states may receive an Autumn 2024 COVID-19 vaccine. Differential equations that define the transitions in the model, and the associated parameter inputs, are provided in the Technical Appendix.

**Figure 1.**
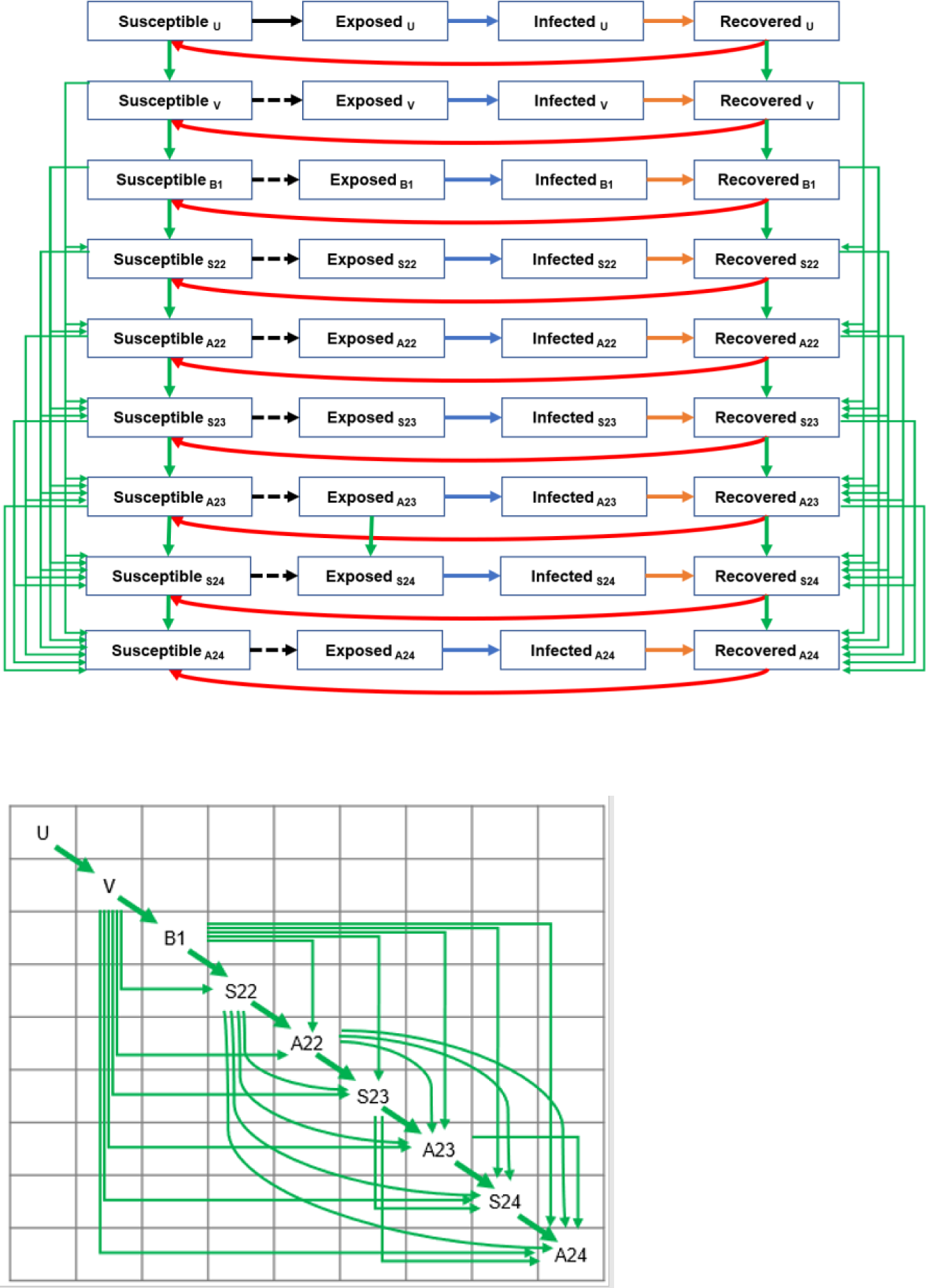
SEIR model structure. Black arrows represent the movement between the susceptible and exposed compartments (force of infection), with dashed black arrows indicating that the force of infection is modified by vaccination compared to the same transition in the unvaccinated stratum. The blue arrows represent the loss of latency which means the infection becomes transmissible. Orange arrows represent the loss of infectiousness which means the infection is cleared and natural immunity develops. Red arrows represent the loss of natural immunity following infection and transition back to the susceptible state. Green arrows represent the possible vaccination points, and the inset box presents more detail on vaccination paths. Please see the Technical Appendix for the mathematical equations driving each transition. Abbreviations: A22, Autumn 2022 booster; A23, Autumn 2023 booster; A24, Autumn 2024 booster; B1, First booster; S22, Spring 2022 booster; S23, Spring 2023 booster; S24, Spring 2024 booster; U, Unvaccinated; V, Primary series vaccination.

Vaccination reduces the incidence of infection (asymptomatic and symptomatic), as well as the risk of hospitalization from a symptomatic infection. Each day, newly vaccinated individuals are assigned the initial VE against infection and hospitalization corresponding to the vaccine used and the variant that is circulating. A waning rate is applied to reduce the VE of previously vaccinated individuals. The average VE for a given day is calculated as a weighted average of the VE for the newly vaccinated individuals and the VE of those vaccinated in past days, weighted by the number of people in each of these groups. The VE against hospitalization (i.e., the reduction in the risk of hospitalization due to infection for vaccinated versus unvaccinated persons) is calculated for use in the infection consequences decision tree (see Technical Appendix).

### Simulation Time Periods

The SEIR model simulation is divided into two time periods: the burn-in period and the analytic time period. The first, the burn-in or model calibration period, begins in January 2020 (the emergence of COVID-19) and runs to August 2024, with a time step of 1 day. This period was used to calibrate the transmissibility parameter and to estimate the residual protection from previously received vaccines and the proportion of individuals with natural immunity at the end of August 2024. In order to calibrate the model, the transmissibility parameter was manually varied over time so that the model predicted the calibration targets, which also varied over time. From January 2020 to October 2022, the calibration targets were the number of infections in the population as estimated by the Institute of Health Metrics and Evaluation (IHME) [27] and the Office of National Statistic (ONS).[28] The calibration target was then switched to be the total number of hospitalizations reported with National Health Service (NHS) England data [28] until January 30, 2024. Please see the Technical Appendix for more detail.

The second is the analytic time period, which is used to evaluate the Autumn 2024 Vaccination Campaign, runs from September 2024 to August 2025 and aligns with the 1-year analytic time horizon for the cost-effectiveness analysis. During this period, assumptions were made about the vaccination effectiveness, vaccine coverage and the transmissibility parameters in order to project the incidence of COVID-19 and the potential public health impact and the incremental cost per quality-adjusted life-years (ICER) of Autumn 2024 vaccines.

For the period of February 2024 to January 2025, which overlaps both the burn-in and analytic time period, the base case transmissibility parameter was assumed to replicate the pattern from February 2023 to January 2024 (See figures in the Technical Appendix). It was then held constant from February 2025 onwards. Three additional incidence scenarios for the analytic time period were created by varying the transmissibility parameter from September 2024 to August 2025. First, an alternative scenario was established in which the transmissibility parameter was held constant over the analytic time horizon with the value based on an average of the last month of the burn-in period. Next, two additional scenarios were created from the base case to model i) increased transmissibility (base case transmissibility was multiplied by a factor of 1.1), and ii) decreased transmissibility (base case transmissibility was multiplied by a factor of 0.9). Finally, a fourth incidence scenario, an immune escape scenario, was created where a new variant was assumed to emerge on April 1, 2025. On this date, it was assumed there was an immediate 40% relative decline in VE (all vaccines; infection and hospitalization outcomes) and natural immunity. Transmissibility was not altered for this analysis.

### Autumn 2024 COVID-19 vaccine effectiveness

The model requires absolute VE inputs as it is structured to compare the vaccinated strata to the unvaccinated model strata. As VE data against hospitalization and infection for absolute effectiveness of the Spikevax XBB.1.5 monovalent vaccine are not yet available, real-world data on the Spikevax bivalent BA.4/BA.5 variant vaccine (mRNA-1273.222) from a prospective study by Tseng et al. (2023)[29] were used. Because the study did not estimate the VE against infection, a meta-analysis by Pratama (2022)[30] which estimated the VE of the original Spikevax monovalent booster (mRNA-1273) against BA1/BA.2 using 9 studies was used. VE input values are summarized in Table 1. Several scenario analyses were conducted by varying the initial VE according to 95% confidence intervals (CIs).

**Table 1.**
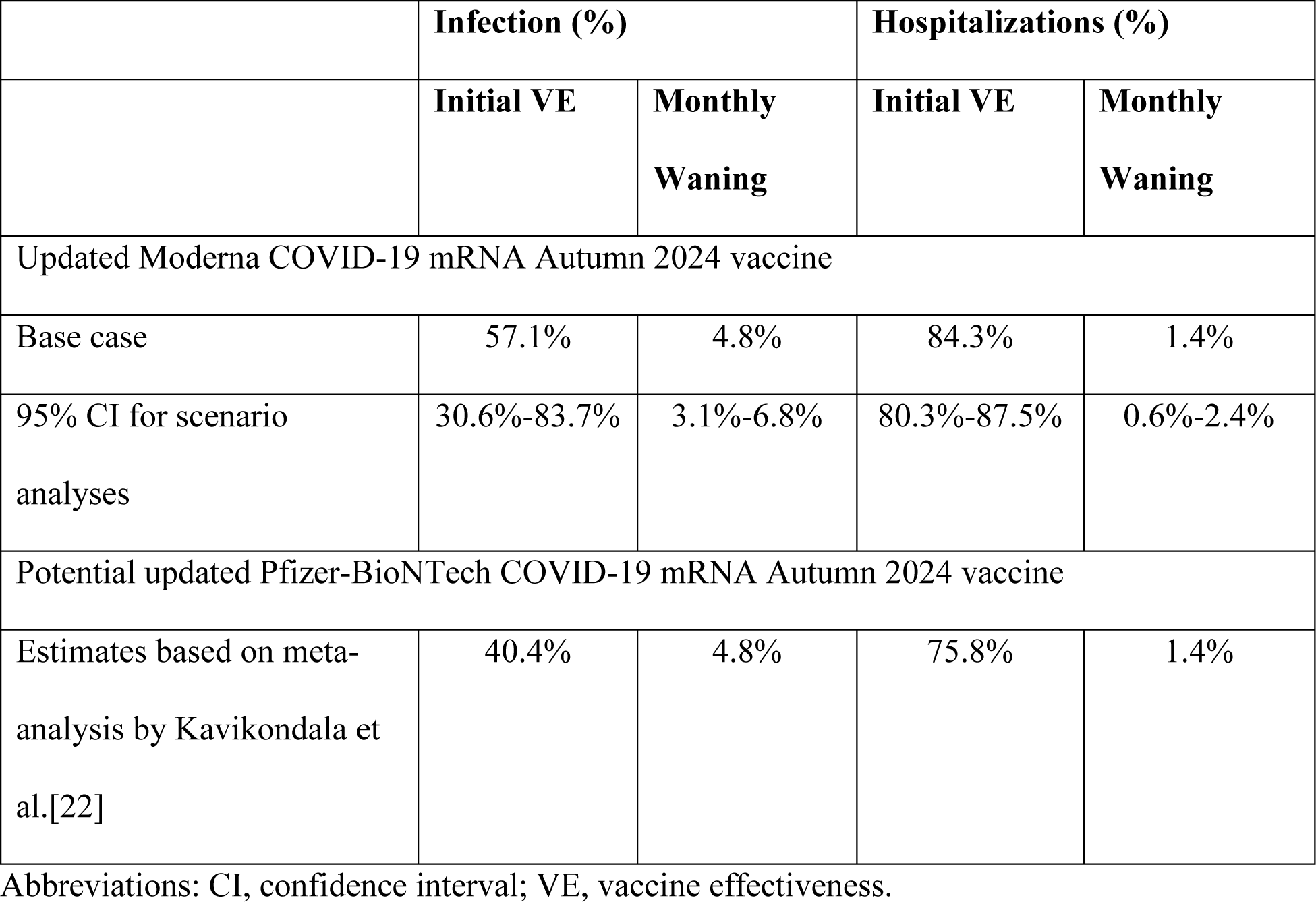
Vaccine effectiveness parameters for the updated COVID-19 mRNA Autumn 2024 vaccines.

|  | **Infection (%)** |  | **Hospitalizations (%)** |  |
| --- | --- | --- | --- | --- |
|  | **Initial VE** | **Monthly Waning** | **Initial VE** | **Monthly Waning** |
| Updated Moderna COVID-19 mRNA Autumn 2024 vaccine |  |  |  |  |
| Base case | 57.1% | 4.8% | 84.3% | 1.4% |
| 95% CI for scenario analyses | 30.6%-83.7% | 3.1%-6.8% | 80.3%-87.5% | 0.6%-2.4% |
| Potential updated Pfizer-BioNTech COVID-19 mRNA Autumn 2024 vaccine |  |  |  |  |
| Estimates based on meta-analysis by Kavikondala et al.[22] | 40.4% | 4.8% | 75.8% | 1.4% |

Abbreviations: CI, confidence interval; VE, vaccine effectiveness.

**Table 2.**
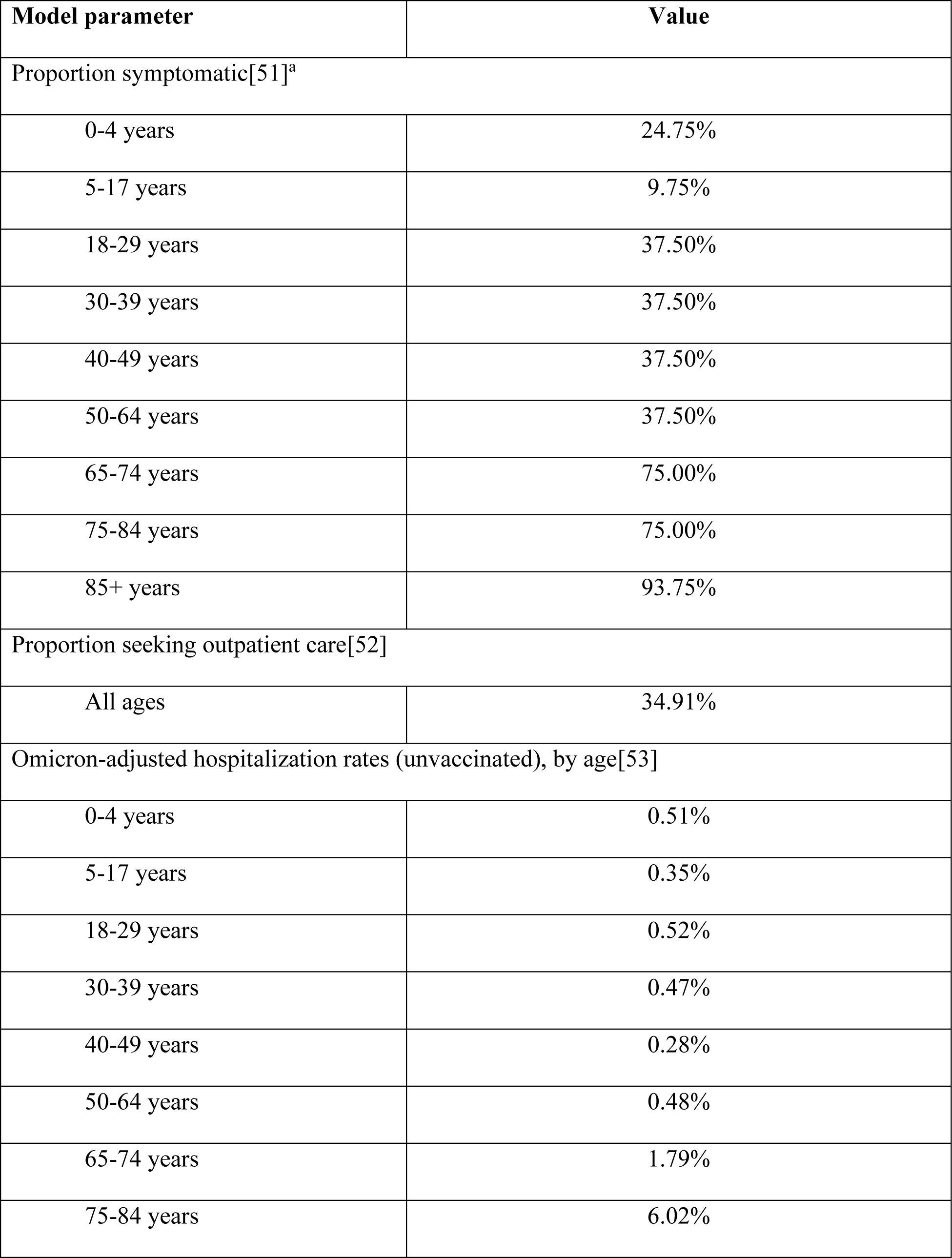
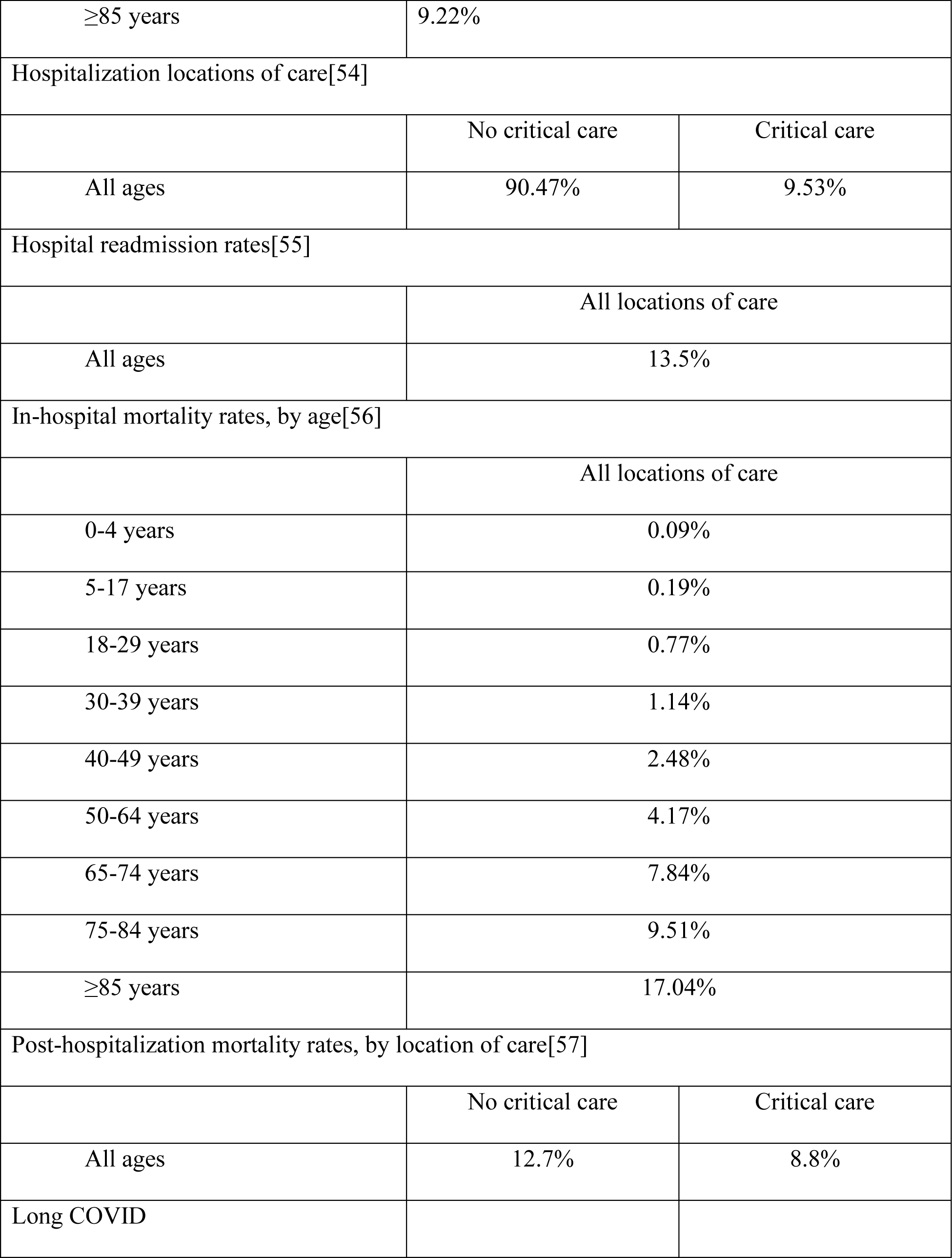
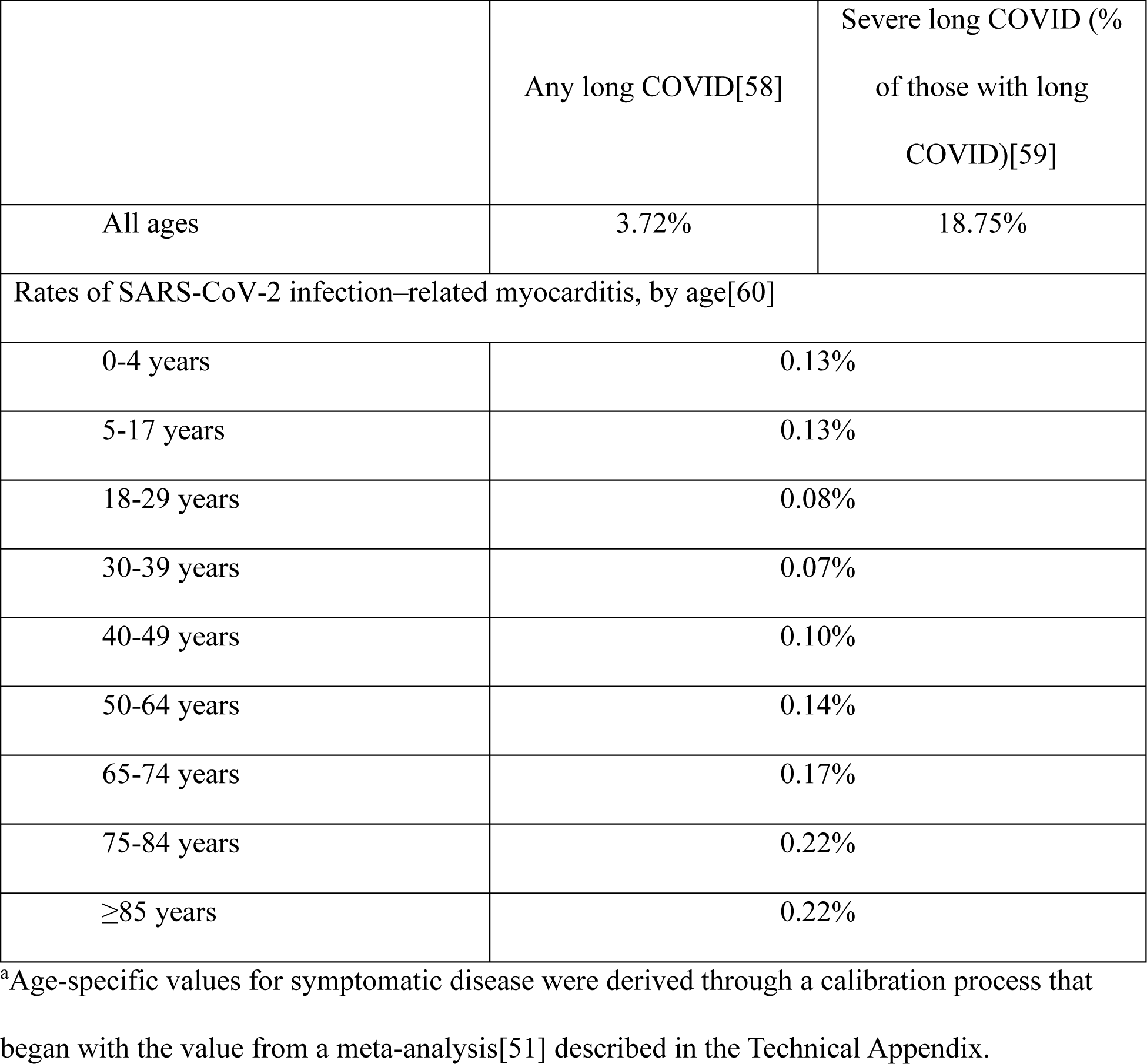
Base case clinical input parameters for the infection consequences model.

| **Model parameter** | **Value** |
| --- | --- |
| Proportion symptomatic[51]<sup>a</sup> |  |
| 0-4 years | 24.75% |
| 5-17 years | 9.75% |
| 18-29 years | 37.50% |
| 30-39 years | 37.50% |
| 40-49 years | 37.50% |
| 50-64 years | 37.50% |
| 65-74 years | 75.00% |
| 75-84 years | 75.00% |
| 85+ years | 93.75% |
| Proportion seeking outpatient care[52] |  |
| All ages | 34.91% |
| Omicron-adjusted hospitalization rates (unvaccinated), by age[53] |  |
| 0-4 years | 0.51% |
| 5-17 years | 0.35% |
| 18-29 years | 0.52% |
| 30-39 years | 0.47% |
| 40-49 years | 0.28% |
| 50-64 years | 0.48% |
| 65-74 years | 1.79% |
| 75-84 years | 6.02% |

|  |  |  |
| --- | --- | --- |
| ≥85 years | 9.22% |  |
| Hospitalization locations of care[54] |  |  |
|  | No critical care | Critical care |
| All ages | 90.47% | 9.53% |
| Hospital readmission rates[55] |  |  |
|  | All locations of care |  |
| All ages | 13.5% |  |
| In-hospital mortality rates, by age[56] |  |  |
|  | All locations of care |  |
| 0-4 years | 0.09% |  |
| 5-17 years | 0.19% |  |
| 18-29 years | 0.77% |  |
| 30-39 years | 1.14% |  |
| 40-49 years | 2.48% |  |
| 50-64 years | 4.17% |  |
| 65-74 years | 7.84% |  |
| 75-84 years | 9.51% |  |
| ≥85 years | 17.04% |  |
| Post-hospitalization mortality rates, by location of care[57] |  |  |
|  | No critical care | Critical care |
| All ages | 12.7% | 8.8% |
| Long COVID |  |  |

|  | Any long COVID[58] | Severe long COVID (%<br>of those with long<br>COVID)[59] |
| --- | --- | --- |
| All ages | 3.72% | 18.75% |
| Rates of SARS-CoV-2 infection–related myocarditis, by age[60] |  |  |
| 0-4 years | 0.13% |  |
| 5-17 years | 0.13% |  |
| 18-29 years | 0.08% |  |
| 30-39 years | 0.07% |  |
| 40-49 years | 0.10% |  |
| 50-64 years | 0.14% |  |
| 65-74 years | 0.17% |  |
| 75-84 years | 0.22% |  |
| ≥85 years | 0.22% |  |

<sup>a</sup>Age-specific values for symptomatic disease were derived through a calibration process that began with the value from a meta-analysis[51] described in the Technical Appendix.

For the comparison of the updated Moderna and Pfizer-BioNTech COVID-19 mRNA vaccines, data from the meta-analysis, which included 24 observational studies, each including two or more doses of Moderna mRNA-1273 or Pfizer-BioNTech BNT162b2 vaccines, was used [22]. This meta-analysis demonstrated a significantly lower risk of infections (including symptomatic and severe infections) and hospitalizations, and a non-significant lower risk of death associated with the mRNA-1273 vaccine compared to the BNT162b2 vaccine in older adults aged ≥50 years. The risk ratios (RRs) for infection 0.72 (95% CI 0.64-0.80) and hospitalization 0.65 (95% CI 0.53-0.79) were used to calculate VEs for the potential updated Pfizer-BioNTech COVID-19 vaccine while the VE of the Moderna vaccine was held constant at the base case value.[22]

The VE of the vaccines were assumed to wane linearly to zero over time. Waning of the future Autumn 2024 boosters (both Moderna and Pfizer-BioNTech) were assumed to be the same as monovalent vaccines against Omicron BA.1/BA.2 and obtained from a meta-analysis by Higdon et al. (2022).[31] Therefore, VE against infection and hospitalization were assumed to decline an absolute 4.8% and 1.4%, respectively, each month. Scenario analyses were conducted for the comparison to no vaccination campaign using the 95% CI (Table 1).

### Autumn 2024 COVID-19 vaccine coverage

Vaccine uptake was assumed to be the same as observed with the 2023 Campaign for the base case analysis.[32] This is appropriate given that the criteria of the Autumn 2023 and Autumn 2024 are very similar. Weekly uptake data from England indicated that most doses were delivered by December 2023 (See Figure 9 in the Technical Appendix). Final coverage was above 70% for those 70 years and older and was minimal for those under 20 years of age (See Table 9 in the Technical Appendix). While household contacts and carers of at-risk individuals are not eligible for the Autumn 2024 vaccine, the coverage data is presented by age only so it is not possible to estimate how many vaccinations were given to these individuals in Autumn 2023. A scenario analysis was conducted where the final cumulative vaccine uptake was decreased by 10%.

A scenario analysis was conducted assuming that the target population is individuals aged 50 years and older and those at high-risk below 50 years of age, meaning that COVID-19 vaccine is offered to a larger cohort than in the Autumn 2023 vaccine campaign. The uptake for the 50-to-64-year age group was based on the Autumn 2022 booster campaign.

### Consequences of infection or vaccination

The SEIR model predicted total number of SARS-CoV-2 infections (asymptomatic and symptomatic) and the incremental reduction in the risk of infection-related hospitalization for vaccinated compared to unvaccinated individuals for each month of the analysis time period. These monthly outputs were then entered into the infection consequences decision tree model to calculate the clinical and economic consequences of infections, which is described in the Technical Appendix.

The expected number of life years lost due to early mortality from COVID-19 was estimated using expected survival by age.[33] Age-specific utility values for individuals without infection[34,35] were applied to the life-year estimates to predict the expected QALYs lost due to early death. All future QALYs lost due to COVID-19 death were discounted by 3.5% annually to present value.[36] Total QALYs lost was calculated as the sum of mortality-related QALYs lost, and QALYs lost due to morbidity, shown in Table 3, and AEs following vaccination. The base case economic analyses were conducted using the healthcare cost perspective. The impact of reducing COVID-19 hospitalizations on the burden to the NHS was assessed by calculating the opportunity cost of bed days required for other hospitalizations using a similar approach to Brassel et al.[37] and Sandmann et al.,[38] where the opportunity cost per COVID-19 hospitalization is the sum of the hospital cost for COVID-19 and the net monetary benefit forgone from the elective hospitalization (additional details are provided in the Technical Appendix). As costs are only counted within the 1-year analytic time horizon, discounting is not required.

**Table 3.** Base case economic and QALY input parameters for the infection consequences model.

| Model parameter | Value | Assumptions |
| --- | --- | --- |
| **Direct costs<sup>a</sup>** |  |  |
| Outpatient care (cost per patient seeking care<sup>b</sup>) | £40.49 | Cost per outpatient visit of £41.13[61] and ED visit of £301.52.[62]<br>0.88[63] outpatient visits and 0.01[63]ED visits. |
| Hospitalization (cost per stay)[64] |  |  |
| No critical care | £2,242.03 | Based on approach by Davis et al.[65]<br>No critical care: weighted average of DX11A and DX21A.<br>Critical care: weighted average of DX01A and DX01A (COVID-related) + XC01Z to XC07Z. |
| Critical care | £24,856.24 |  |
| Hospitalization recovery (cost per case) | £329.60 | Cost per chest x-ray of £41.41;[66] cost of general practitioner e-consultation of £41.13;[61] 2.0[67] chest x-rays and 6.0[67] general practitioner e-consultations. |
| Infection-related myocarditis |  |  |
| 0-17 years | £8052.58 | Weighted average over codes EB14A-E.[64] |
| 18+ years | £3972.45 |  |
| Post-infection costs, by age and hospitalization status |  |  |
| All ages | £14.22 | Estimated annual cost beyond week 12 post-acute infection period in non-hospitalized individuals. Assumed same cost of hospitalized due to lack of data.[52]<br>Values inflated using the NHS Cost Inflation Index.[61] |
| Long COVID | All locations of care |  |
| Long COVID - All ages | £67.69 | Annual primary care costs for non-hospitalized individuals.[52] No data available for hospitalized individuals, and therefore assumed same value. |
| Severe long COVID – All ages | £2,193.46 | ICD code for long COVID (U074) mapped to HRG codes WH15 and PX57.[64,68] |
| SARS-CoV-2 infection–related myocarditis (cost per event), by age |  |  |
| 0-17 years | £8052.58 | Applied to all patients with infection-induced myocarditis (assumes 100% hospitalized). |
| 18+ years | £3972.45 |  |
| QALY decrements |  |  |
| Infection-related myocarditis[69] | 0.0038 |  |
| Symptomatic infection, not hospitalized[70] | 0.0043 | Applied whether or not medical care was sought. |
| Hospitalized, not critical care[71] | 0.0168 | The symptomatic infection, not hospitalized decrement is also added to these values to reflect symptoms prior to admission. |
| Hospitalized, critical care[71] | 0.033 |  |
| Hospital readmission[70,71] | 0.029 | Average of non-critical care and critical care, with the addition of symptomatic infection, not hospitalized. |
| Post-infection, not hospitalized[72] | 0.024 | Based on data measured at 6 months following initial infection. |
| Post-infection, hospitalized[73] | 0.091 | Based on follow-up data with a mean of 48 days post-discharge from hospital. |
| **Opportunity cost and bed day parameters** |  |  |
| Hospitalization cost / LOS: COVID-19[64] | £4397.16 / 8.96 days | Weighted cost of no critical care and critical care hospitalizations. |
| Hospitalization cost / LOS: elective inpatient[64] | £5939.93 / 5.10 days | NHS cost collection database. To calculate, all COVID-related cost codes and all codes related to the pediatric population were excluded. The average of all remaining cost codes was then calculated. |
| QALYs gained for a non-COVID treatment[37] | 0.2390 |  |
| Willingness-to-pay threshold | £20,000 | JCVI |

Abbreviations: ED, emergency department; HRG, Health Resource Group; ICD, International Statistical Classification of Diseases; JCVI, Joint Committee on Vaccination and Immunisation; LOS, length of stay; QALY, quality-adjusted life-year; NHS, National Health Service.
<sup>a</sup>Costs are reported in 2023 GBP.
<sup>b</sup>Cost inputs for outpatient care are weighted by the proportion of patients who seek medical care, with a cost of zero assigned to those who do not seek care.

Separately, the consequences of vaccination were also estimated. The costs and QALY decrements associated with vaccine-related AEs, including Grade 3 local and systemic solicited events, anaphylaxis, and vaccine induced myocarditis/pericarditis, were calculated as described in the Technical Appendix. The unit cost of the updated Moderna Autumn 2024 vaccine was set to a proxy price of £67, while the unit cost of the updated Pfizer-BioNTech Autumn 2024 vaccine was set to £64.[39] A vaccine administration cost of £7.54 was also included.[40]

### Scenario and deterministic sensitivity analyses (DSAs)

A series of deterministic sensitivity analyses (DSAs) were conducted to test the robustness of the base case analysis results. Several of these have been described above in the text and include SEIR parameters related to infection incidence, percentage of infections that are symptomatic, Autumn 2024 vaccine coverage and VE. In addition, the infection consequences decision tree parameters were varied. Where available, rates were varied according to their 95% CIs. For all other DSAs, parameters were varied by ±25% of their base case value. As the Moderna Autumn 2024 vaccine unit cost was not finalized, an analysis where the unit cost was varied until the incremental cost-effectiveness was equal to the willingness-to-pay threshold of £20,000/QALYs gained was conducted.

Additionally, several scenario analyses for the comparison to no vaccination campaign were conducted: i) vaccination of all persons aged ≥50 years plus at-risk individuals <50 years; ii) use of alternate perspectives (healthcare system cost perspective excluding costs associated with opportunity cost for bed days; and societal perspective including productivity losses [see Technical Appendix for related input parameters]); iii) vaccination of all persons aged ≥50 years plus at-risk individuals <50 years using the healthcare and societal perspectives; iv) use of parameters from a previous analysis by Shiri and colleagues (one analysis utilizing their hospitalization rates and a second using their hospital mortality rate),[41] and v) not discounting the QALYs lost associated with early death due to COVID-19.

For the comparison to the Pfizer-BioNTech Autumn 2024 vaccine, an additional analysis was conducted where its vaccine coverage was systematically increased until the total healthcare treatment costs were equivalent to those predicted when the Moderna Autumn 2024 vaccine was used.

## Results

### Comparison: Moderna Autumn 2024 Campaign versus No Autumn 2024 Campaign

For the base case analysis, the model predicted that from September 1, 2024, to August 31, 2025, there will be 8,257,540 symptomatic infections without an Autumn 2024 COVID-19 vaccine campaign compared to 6,674,735 symptomatic infections if 12,796,178 vaccines are administered during the Moderna Autumn 2024 Campaign (Table 4), representing a decrease of 1,582,805 symptomatic infections (−19%). The predicted number of daily infections over time is shown in Technical Appendix Figure 13. At the start of the analytic time period, only those who received a booster in Autumn 2023 or Spring 2024 have residual protection against infection. As these individuals are primarily in the older age groups (those ages ≥65 years), it can be noted that the rate of infection surges during the analytic time period as many of the younger age groups who spread the infection no longer have vaccine-mediated immunity.

**Table 4.**
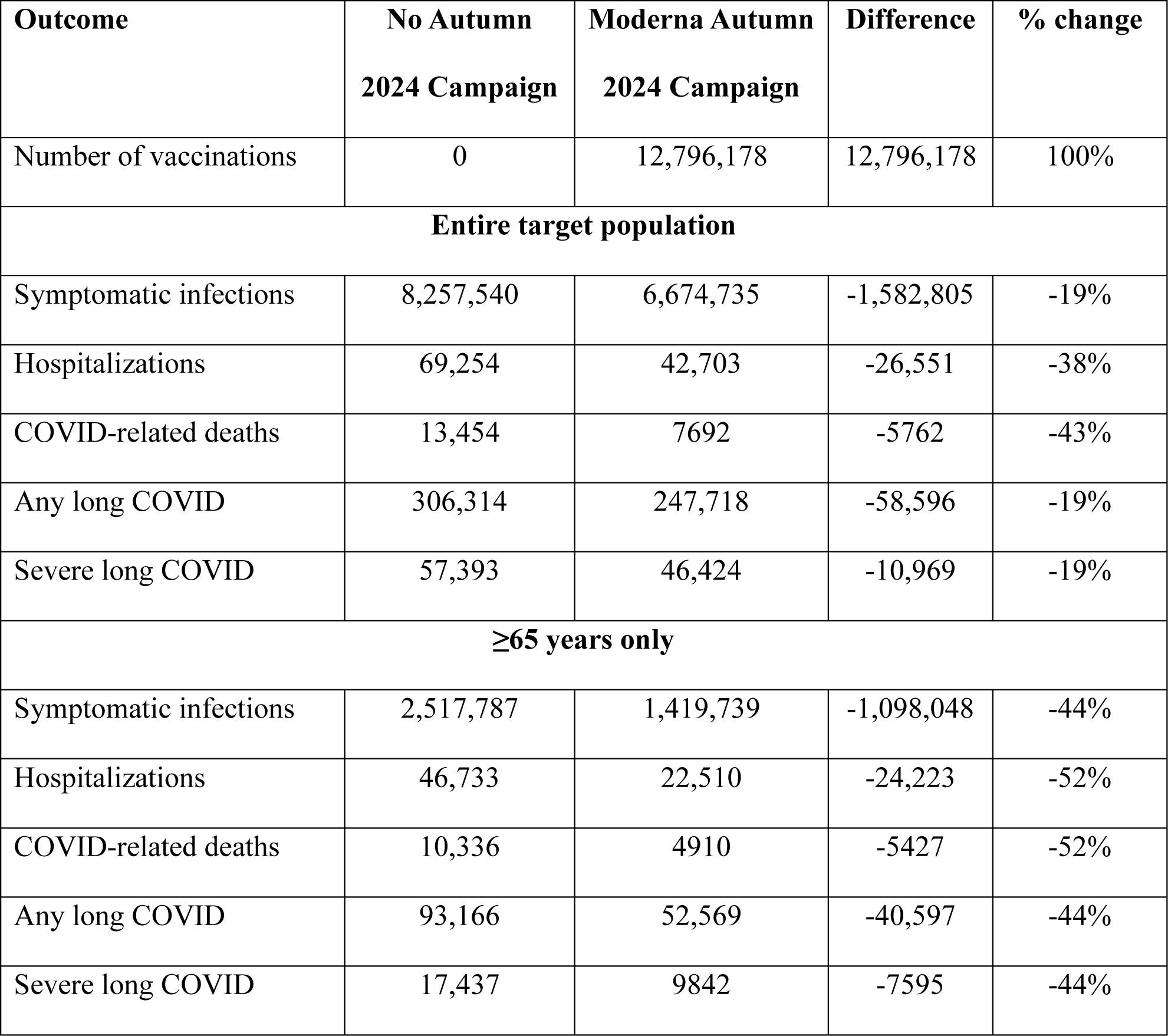
Clinical outcomes with and without the Moderna Autumn 2024 Campaign.

While infections are predicted to be higher during the analytic time horizon than in the previous year, the number of hospitalizations is not. At the start of the analytic time horizon, there is still at least 40% VE against hospitalization in all individuals who received the Autumn 2022 booster and at least 60% VE against hospitalization in those who received vaccines during or after the Spring 2023 booster campaign. Between September 2022 and August 2023, there were 67,662 reported hospitalizations in the full population and 46,348 reported hospitalizations among those ≥65 years.[42] Between February 2023 and January 2024, these counts had dropped to 53,047 and 36,039, respectively. For the base case, the model predicted 46,733 hospitalizations from September 2024 to August 2025 in those ≥65 years when there was No Autumn 2024 vaccine Campaign and 22,510 with the Moderna Autumn 2024 Campaign (Table 4). Therefore, despite a surge in infections, the predicted hospitalizations are lower than previous years when the Moderna Autumn 2024 Campaign is implemented. The counts of other COVID-19 related outcomes and the difference with vaccination are also presented in Table 4, both for the entire target population as well as for the proportion of the target population that is ≥65 years. This stratification highlights that the majority of hospitalizations (67%) and deaths (77%) occur in those who are ≥65 years; hence, targeting older age groups for vaccination decreases overall hospitalizations and deaths in the population by 38% and 43%. Given only 30% of infections, long COVID, and severe long COVID occur in this population, the resultant reduction of these outcomes is 19% only in the base case analysis.

Given this clinical impact, the Moderna Autumn 2024 Campaign is expected to lead to a gain of 32,532 QALYs by preventing COVID-19–related deaths and 45,392 QALYs gained due to prevented morbidity, totaling 77,925 QALYs gained than without an autumn 2024 campaign. Vaccination costs £954 million; however, COVID-19 treatment costs with vaccination are £700 million compared to £989 million without the vaccine, leading to £665 million in costs prevented with vaccination. The incremental cost per QALY gained for the Moderna Autumn 2024 Campaign compared to No Autumn 2024 Campaign is therefore £8540 (Table 5; see Technical Appendix for further QALY and cost disaggregation). Considering a threshold of £20,000 per QALY gained, the unit cost of the Moderna Autumn 2024 vaccine can be increased to £136.79 and still be considered cost-effective compared to no vaccination campaign under base case conditions.

**Table 5.**
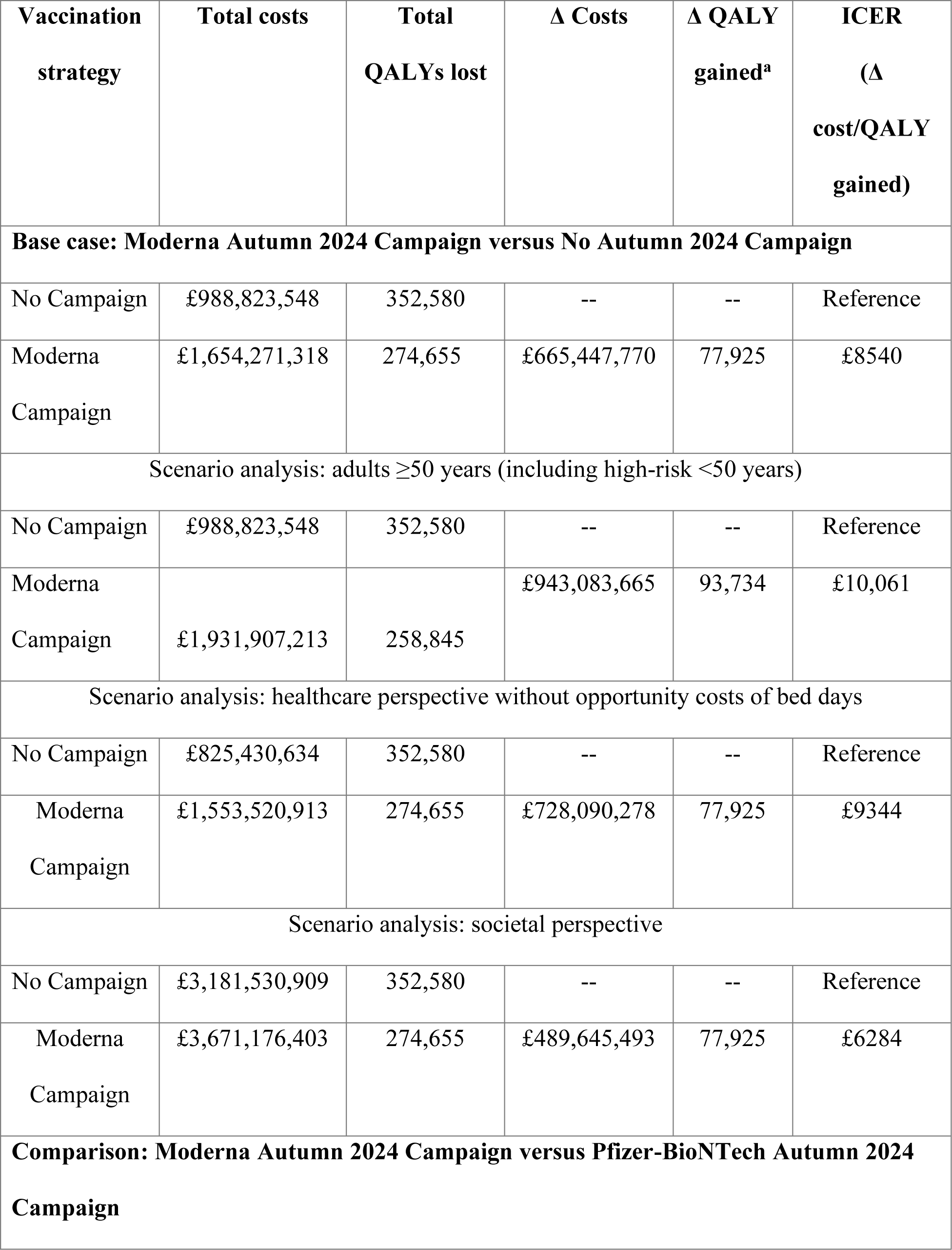
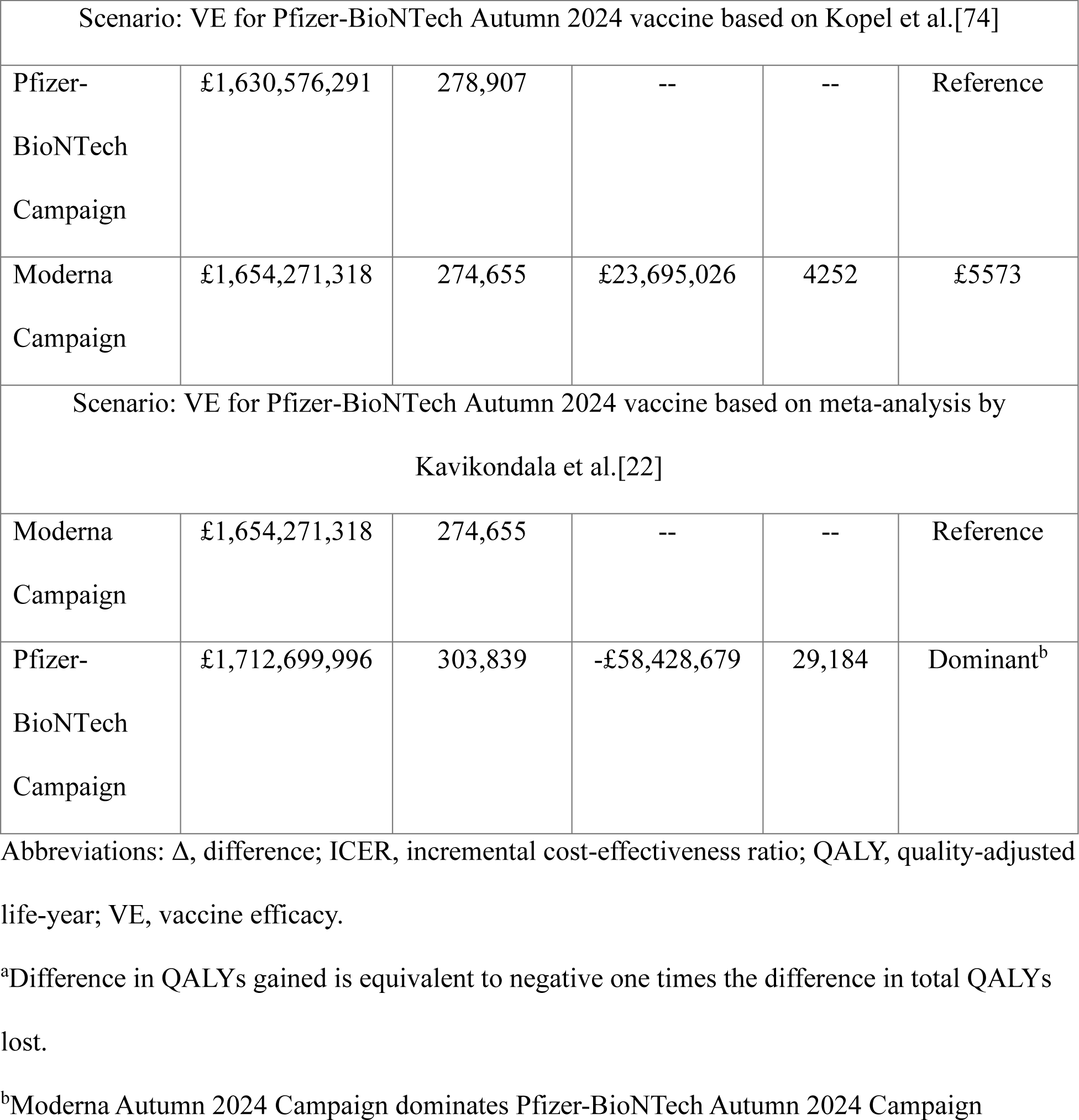
Economic results for the base case and key scenario analyses.

When the size of the target population was increased by opening up vaccination to all persons aged ≥50 years (including those at at-risk ˂50 years), the clinical impact of the Moderna Autum 2024 Campaign increases compared to No Autumn 2024 Campaign. Overall, there is an increase of 4,295,002 vaccines (34%) compared to the number administered within base case target population. However, there are 378,850 (6%) more infections, 2952 (7%) more hospitalizations, 550 (7%) more deaths and 13,603 (6%) more severe long COVID cases prevented compared to the base case target population. Increasing the number of vaccines provided increases the total cost for the Moderna Autumn 2024 Campaign strategy, but the strategy remains cost-effective with an ICER of £10,061/QALY gained.

When the cost perspective is changed in scenario analyses, the Moderna Autumn 2024 Campaign remained cost-effective (See Table 5 for full results). If the opportunity costs of bed days is excluded from the healthcare perspective, the ICER increases to £9344 per QALY gained. With the societal perspective that includes productivity loss, the ICER decreases to £6284 per QALY gained. Considering the expanded population that includes all persons aged ≥50 years, the ICER is £7,606.

The results of the key DSAs are displayed in the tornado diagram in Figure 2 and a table with all DSAs is provided in the Technical Appendix. Varying the initial VE of the Moderna Autumn 2024 vaccine has the most impact on the ICER. With the low VE values, 7% of symptomatic infections and 27% of hospitalizations are prevented compared to 31% and 49% with the high VE values. The next most sensitive inputs are the rate of Moderna Autumn 2024 VE waning over time, hospitalization rates in the unvaccinated, proportion of infections that are symptomatic, post-infection QALY loss, hospital mortality rate, and infection incidence (Figure 2). All of the remaining parameters change the ICER by less than 10% compared to base case results.

**Figure 2.**
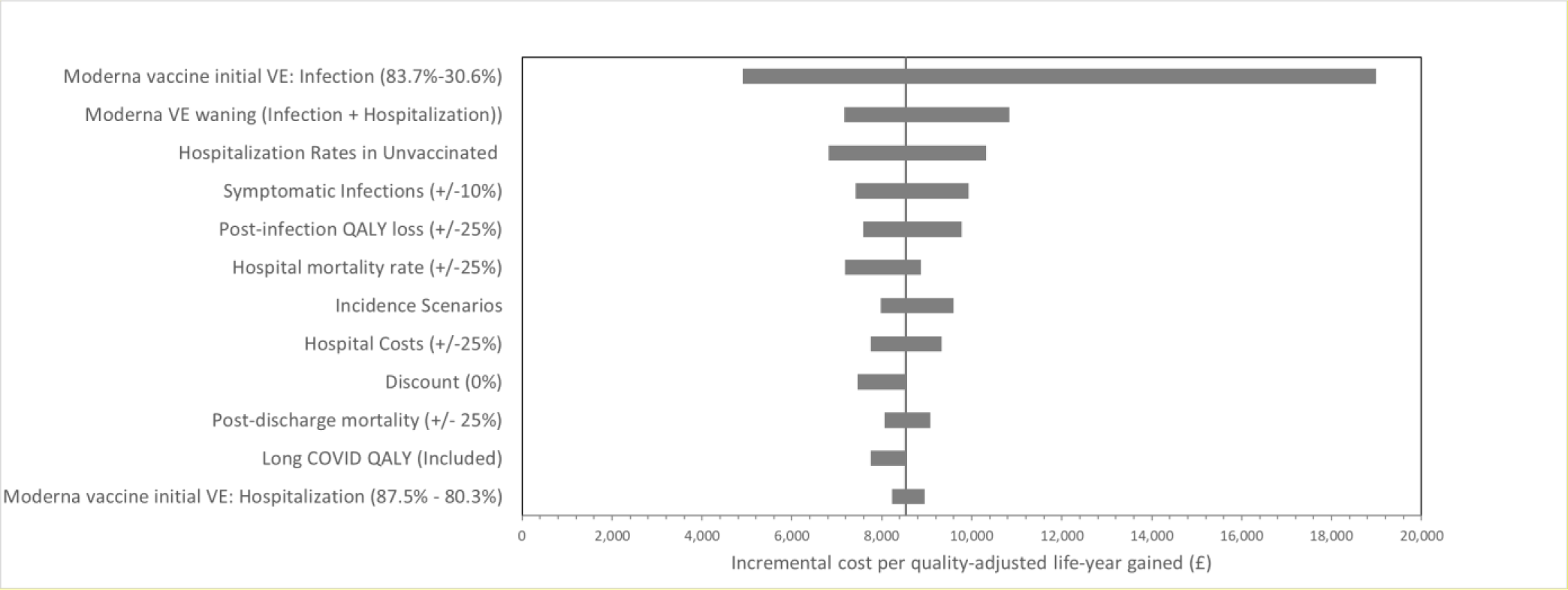
Tornado diagram: key deterministic sensitivity analyses. Abbreviations: QALY, quality-adjusted life-year; VE, vaccine effectiveness.

### Comparison: Moderna Autumn 2024 Campaign versus Pfizer-BioNTech Autumn 2024 Campaign

Given the expected lower VE of the potential updated Pfizer-BioNTech Autumn 2024 vaccine estimated using data from a meta-analysis in which data from primary series and first booster campaigns were aggregated,[22] use of the Moderna Autumn 2024 vaccine is predicted to prevent 645,165 more SARS-CoV-2 infections than the potential Pfizer-BioNTech Autumn 2024 Campaign over the 1-year time horizon. In addition, use of the Moderna vaccine is predicted to result in 7920 fewer hospitalisations and 1696 fewer deaths than use of the potential Pfizer-BioNTech vaccine. This is expected to lead to an additional 29,184 QALYs gained. Despite the higher cost of the Moderna vaccine, savings from the clinical events avoided leads to cost savings that fully offset its higher unit cost and, hence, the Moderna Autumn 2024 Campaign is predicted to dominate the Pfizer-BioNTech Autumn 2024 Campaign (Table 5; additional results are provided in the Technical Appendix).

When vaccine coverage was the same for Moderna and Pfizer-BioNTech, healthcare costs were £78 million higher with Pfizer-BioNTech due to its lower VE. In order to decrease the health care treatment costs (e.g. the costs that are not associated with the vaccine), the coverage of the Pfizer-BioNTech was systematically increased for all ages. When the Pfizer BioNTech vaccine coverage was increased by 15% across all age groups (Figure 3), meaning that 8,873,115 additional vaccines were delivered, there was only a £1 million difference in health care treatment costs between Moderna and Pfizer-BioNTech (see Technical Appendix Table 17 for detailed clinical outcomes) implying that coverage with a Pfizer-BioNTech vaccine would need to be increased by 15% to achieve similar savings in healthcare treatment costs as compared to a campaign with a Moderna vaccine.

**Figure 3.**
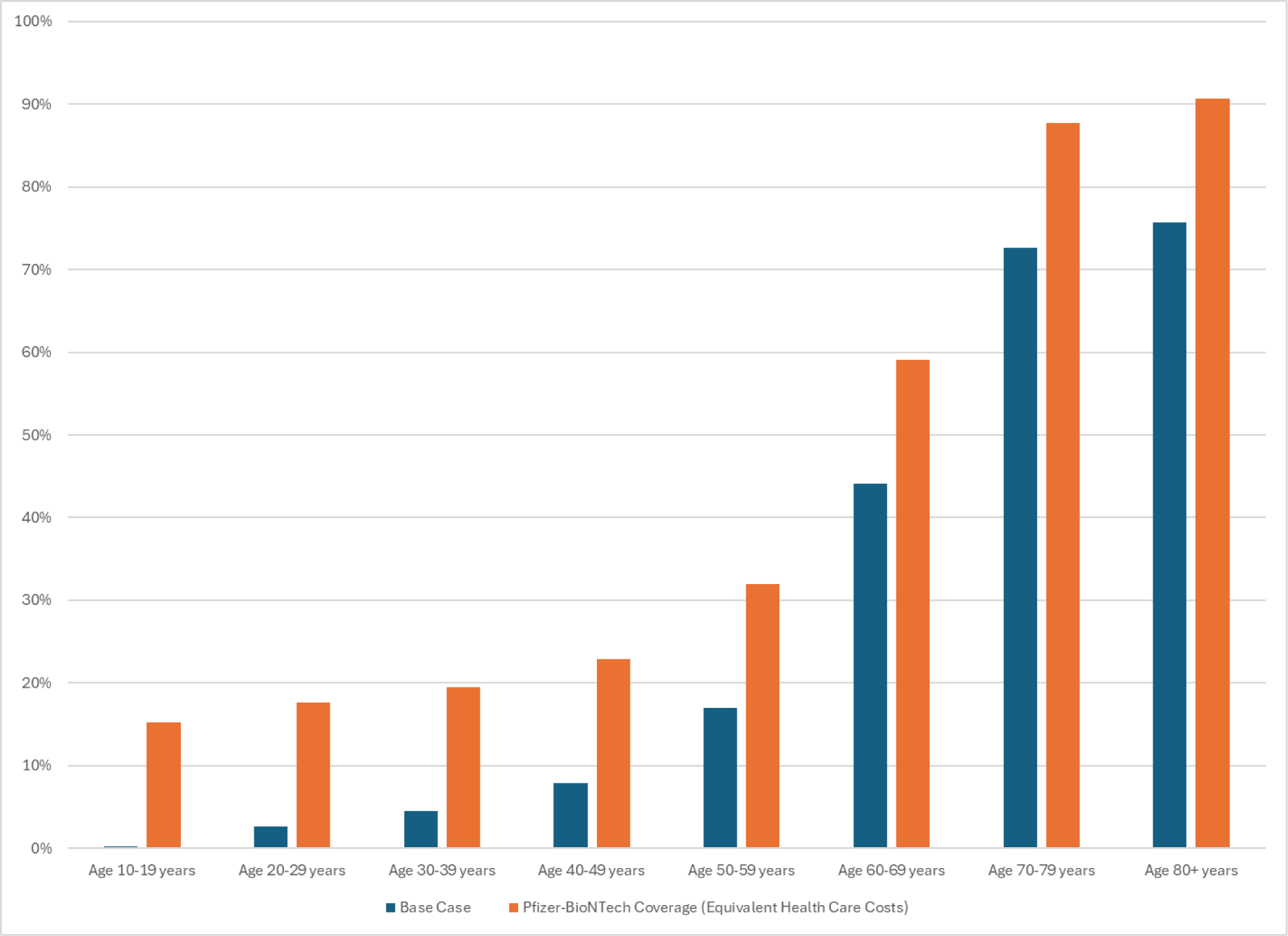
Base case and increased coverage of the Pfizer-BioNTech vaccine. With increased coverage scenario the difference in healthcare costs (without vaccine costs) from the Moderna Autunm 2024 scenario is only £1 million.

## Discussion

In the UK, a proactive, evidence-led vaccination approach is required to effectively manage COVID-19, particularly given the uncertainty surrounding the nature of evolving virus variants. The national vaccination effort, guided by the JCVI, focuses on safeguarding the most vulnerable and the elderly, groups at risk of higher COVID-19 related hospital admissions. In anticipation of a “complex Autumn season”, NHS England is implementing an autumn/winter plan with the objective of improving the uptake of all seasonal vaccinations, including COVID-19 doses.[43] Amid the continued presence of the SARS-CoV-2 virus, a vaccination strategy that is both sensible and cost-effective is of utmost importance.

In this analysis, the potential clinical and economic impact of an Autumn 2024 vaccine campaign using an updated Moderna COVID-19 mRNA vaccine for those ≥65 years of age and those ages 6 months to 64 years at high-risk of complications of COVID-19 infections was determined. The strategy was predicted to be highly cost-effective, with a base case incremental cost per QALY gained of £8540 compared to No Autumn 2024 Campaign. The vaccine would still remain cost effective at a WTP of £20,000 if doubled in price.

The vaccine campaign is predicted to have a higher impact on hospitalizations than on infections because of the characteristics of the eligible population. Most of the vaccines are received by those aged 65 years and older who experience higher rates of severe disease with a SARS-CoV-2 infection. As a side note, one of the limitations of this analysis is that the SEIR model only tracks individuals by age and not by risk of severe outcomes. While vaccines are targeted to at-risk individuals in those under 65 years of age because of poorer outcomes following infection, the model can only assign an average risk of hospitalization by age group. Therefore, the model may under-estimate the expected decrease in hospitalizations amongst in those vaccinated at-risk persons aged less than 65 years. In the simulations, the majority of infections occur in younger age groups because they have more frequent daily contacts[44] and are more likely to spread respiratory infections than older age groups. As only a small portion of the age groups responsible for spreading infection are expected to receive the vaccine under the Autumn 2024 campaign, less than 20% of infections in the population are predicted to be prevented.

As with all decision-analytic studies, this analysis is subject to limitations related to the uncertainty of the data. DSAs conducted to determine the potential impact of that uncertainty demonstrated that a key driver of the cost-effectiveness of vaccination is the initial VE achieved in the first month following vaccination. The range of the VE values used are based on 95% Cis from the source study, which examined the bivalent vaccine.[29] Much of the recently published real-world data on the updated XBB vaccine are from studies comparing individuals who have received updated vaccines to previously vaccinated individuals who did not receive an updated vaccine and therefore are estimates of relative VE (rVE). Given that VE wanes over time, the rVE will depend on the time since previously received vaccination, making it difficult to directly compare the rVE from various published studies to absolute VE.

Early data from a US study provides an estimate of the VE of the monovalent XBB.1.5 vaccines, including versions from both Moderna and Pfizer-BioNTech, against infection.[45] A VE of 58% was estimated for those who received an XBB1.5 vaccine 7 to 59 days earlier relative to those that did not receive a dose. The VE was similar for the sub-group who were either unvaccinated or received an original monovalent dose a median of 765 days earlier (57%) and the sub-group who received a bivalent dose a median of 365 days earlier (56%). As the VE appears to be consistent when comparing to those who received a COVID-19 vaccine one year ago or two years ago, it is plausible that the rVE compared to a dose given one year prior is consistent with estimates of absolute VE.

Preliminary data on VE against hospitalization for XBB.1.5 vaccines (not specific to Moderna) are available from a Danish study. Hansen et al. estimated a hazard ratio (HR) of 0.239 for hospitalizations for those who had received the XBB.1.5 vaccine compared to those that only received the bivalent vaccine, translating to a rVE of 76.1%.[46] Assuming 12 months since last vaccination in the controls, giving an absolute VE remaining of 66.6%, the absolute VE for the XBB dose is 92.0%. This is higher than the VE assumed for the Autumn 2024 vaccine in the base case and the scenario analyses suggesting a conservative approach was taken in our analyses. Similarly, the interim results from the Moderna study of the XBB.1.5 vaccine estimated the rVE against hospitalization at 60.2%. [47] Assuming that most in the control group received their last vaccine 12 months ago, the absolute VE would 86.7%. Once again, this absolute VE is within the range of the scenario analyses already conducted.

Given the uncertainty still associated with the future of COVID-19 incidence, the Department of Health and Social Care conducted an impact analysis[48] varying the risk of infection using high, low, and most plausible scenarios. Consequently, uncertainty in COVID-19 incidence was evaluated in our model through various scenarios, demonstrating robustness with ICER values ranging from £7976 to £9598. Incidence was an input to the Department of Health and Social Care (DHSC) analysis as the model itself could not generate estimates of incidence. For this analysis, the transmissibility pattern from the past year was used by our dynamic model to project future COVID-19 incidence. Given the model design, it was not possible to set incidence equivalent to the past year. Even with the same transmissibility pattern, projected incidence may not be the same as the previous year due to changing patterns of natural and vaccine-mediated immunity. However, for the base case scenario, the predicted number of hospitalizations were lower than in the past year when an Autumn 2024 booster is given and therefore, we believe this is a plausible future scenario.

Sandmann and colleagues also assessed the potential economic benefit of COVID-19 vaccines in the UK by using a dynamic model to explore potential future incidence scenarios.[49] However, their analysis was conducted during the pandemic, prior to the emergence of the Omicron variant. Since then, population immunity has increased and COVID-19 incidence has therefore declined.

As one of the stated goals of the JCVI is to reduce the burden of COVID-19 on the NHS, especially during the winter months when other respiratory infections are expected to peak, the additional benefit of reducing COVID-19 hospitalizations is included by calculating the opportunity cost of bed days required for other hospitalizations. The potential impact of these costs was also considered in the Impact Analysis conducted by the Department of Health for the 2023 Autumn vaccine.[48] They stated that these were “higher than usual pressures on healthcare resources” as the NHS recovers from the COVID-19 pandemic. When health system stress costs are excluded, the ICER increases to £9344 but remains cost-effective.

The societal perspective included only patient productivity loss and did not include estimates of caregiver time productivity loss. With the societal perspective, the ICER decreased by £2256 to £6284 per QALY gained.

When the Autumn 2024 vaccine is offered to an expanded cohort that includes all adults ages ≥50 years, SARS-CoV-2 infections, COVID-19 related hospitalizations and deaths, and cases of long COVID are predicted to be reduced by a further 6 to 7% while remaining cost-effective at a WTP threshold of £20,000 per QALY gained. This reduction shows the importance of considering the value of a broad vaccination program to strengthen the healthcare system, especially during the winter season. Including the societal perspective drops the ICER by £2455 to £7606.

While protection against infection is not as durable as protection against hospitalization, vaccination of younger cohorts has value in reducing the level of circulating virus during the winter season. The SEIR model predicts that reduction in infection in younger, lower risk age groups will indirectly benefit the higher risk populations by reducing their infections as well. This effect would further reduce the associated hospitalizations in older adults at a time when the NHS may be experiencing considerable added strain due to increases in other respiratory infections.

In the analysis, long COVID is assumed to affect all adults equally and therefore vaccination of younger age groups reduces this post-infection outcome. ONS data indicate that younger individuals are more likely to experience long COVID in the UK.[50] Therefore, our analysis likely provides an underestimate of the true impact of COVID-19 in this age group.

The analysis comparing the Moderna and Pfizer-BioNTech Vaccination Campaigns found additional cases of infections, hospitalizations and deaths prevented due Moderna vaccine’s predicted higher VEs, resulting in health cost savings. Even with increasing the Pfizer-BioNTech coverage by 15% among all age groups, there is still a £1 million cost savings with the use of the Moderna vaccine. Quantifying the difference in vaccine choices is important given the upcoming tendering environment in 2025.

## Conclusions

Vaccination in the ≥65 and high-risk population ˂65 years of age contributes to significant reductions in hospitalizations, death and long COVID compared to no further vaccination. Additionally, expanding the eligible population to those ≥50 years of age continues to remain cost-effective. If the difference in VE between the two Autumn 2024 mRNA vaccines is similar to that currently reported in the literature for past versions, then the Moderna vaccine will be cost-effective (or possibly cost-saving) compared the Pfizer-BioNTech vaccine, despite a higher unit cost. Hence, use of the Moderna Autumn 2024 Campaign is predicted to reduce COVID-19 infections and associated outcomes in a cost-effective manner and will contribute to a more resilient healthcare system in the UK.

## Supporting information

Technical appendix

## Data Availability

All data produced in the present work are contained in the manuscript

## Transparency

### Author contributions

AL, EB, KJ, MK, MM, NV, OB and SC were involved in study design and interpretation of the analysis. MM programmed the model with quality assurance by MK and AL. All authors were involved in model estimation. MM and MK conducted the analysis. AL, MK, MM and OB wrote the initial draft of the manuscript, and all remaining co-authors critically revised the manuscript and approved the final version.

## Acknowledgements

We acknowledge Heather Davies and Sarah Medland from the York Health Economics Consortium (YHEC) for their support on the derivation of input parameters for this study.

