## Supplementary material for "The potential clinical impact and cost-effectiveness of the updated COVID-19 mRNA Autumn 2024 vaccines in the United Kingdom": Technical appendix

**Tables**

**Figures**

1. SEIR model inputs: burn-in period (January 2020 to August 2024)

The model structure and underlying assumptions of the Susceptible-Exposed-Infected-Recovered (SEIR) model were described in the Technical Appendix of the previously published analyses.[1,2] United Kingdom (UK) specific data and methods are outlined in the main manuscript and below.

### 1.1. Number of susceptibles

All individuals in the UK were included in the model and were considered to be susceptible to SARS-CoV-2 infection at the start of the SEIR model simulation (January 2020). The size of the UK population in 2020, by age, was obtained from the the Office for National Statistics (ONS).[3]

Table 1. Model population size

| **Age group (years)** | **Number in population** |
| --- | --- |
| 0 to 4 | 3,782,330 |
| 5 to 9 | 4,147,413 |
| 10 to 14 | 4,045,114 |
| 15 to 19 | 3,683,680 |
| 20 to 24 | 4,133,158 |
| 25 to 29 | 4,476,630 |
| 30 to 34 | 4,521,975 |
| 35 to 39 | 4,404,100 |
| 40 to 44 | 4,091,543 |
| 45 to 49 | 4,303,967 |
| 50 to 54 | 4,616,017 |
| 55 to 59 | 4,510,851 |
| 60 to 64 | 3,855,818 |
| 65 to 69 | 3,355,381 |
| 70 to 74 | 3,363,906 |
| 75 to 79 | 2,403,759 |
| 80 to 84 | 1,726,223 |
| 85 and over | 1,659,369 |
| **Total** | **67,081,234** |

### 1.2. Number of effective contacts

Contact matrices were used to determine age-specific mixing patterns. These matrices are modified by a mobility index (accounting for social distancing and mask use) as behaviors changed over time during the pandemic.

#### 1.2.1. Mixing patterns/contact matrices

Data on the age-specific mixing patterns in the general population for the UK were obtained from Prem and colleagues.[4] Details on these contact matrices and their application are provided in the Technical Appendix of the US analysis by Kohli et al.[2]

The base contact matrix is presented in Table 2.

Table 2. Base contact matrix used in the model before applying scaling factors

| **Age group of**  **participant (years)** | **Age group of contact (years)** | | | | | | | | |
| --- | --- | --- | --- | --- | --- | --- | --- | --- | --- |
|  | **0-9** | **10-19** | **20-29** | **30-39** | **40-49** | **50-59** | **60-69** | **70-79** | **80+** |
| **0-9** | 3.67 | 1.20 | 0.80 | 1.44 | 0.73 | 0.30 | 0.23 | 0.08 | 0.03 |
| **10-19** | 1.17 | 7.71 | 1.28 | 1.15 | 1.60 | 0.61 | 0.23 | 0.35 | 0.30 |
| **20-29** | 0.87 | 1.43 | 3.92 | 2.00 | 1.86 | 1.38 | 0.53 | 0.42 | 0.18 |
| **30-39** | 1.62 | 1.33 | 2.07 | 3.25 | 2.25 | 1.37 | 0.76 | 0.35 | 0.54 |
| **40-49** | 0.77 | 1.74 | 1.81 | 2.11 | 3.33 | 1.55 | 0.70 | 0.76 | 0.95 |
| **50-59** | 0.35 | 0.72 | 1.46 | 1.40 | 1.68 | 1.87 | 0.91 | 0.60 | 0.65 |
| **60-69** | 0.21 | 0.21 | 0.44 | 0.61 | 0.60 | 0.72 | 1.01 | 0.66 | 0.60 |
| **70-79** | 0.05 | 0.26 | 0.28 | 0.22 | 0.52 | 0.38 | 0.53 | 0.86 | 1.36 |
| **80+** | 0.01 | 0.13 | 0.07 | 0.20 | 0.38 | 0.24 | 0.28 | 0.80 | 1.48 |

Note: Since the population size of the age groups are not of equal size, the transformed contact matrix is not strictly symmetric.

#### 1.2.2. Social distancing and mask use: overall scaling factor

The rate of contact during the pandemic was reduced by behaviors such as social distancing and mask use. Methods are described in the Technical Appendix of the US analysis by Kohli et al.[2]

The final monthly pattern for the scaling factor for both the burn-in period (January 2020 – August 2024) and the analysis period (September 2024 – August 2025) is shown in Figure 1.

Figure 1. Mobility scaling factor over time (burn-in and analysis period)

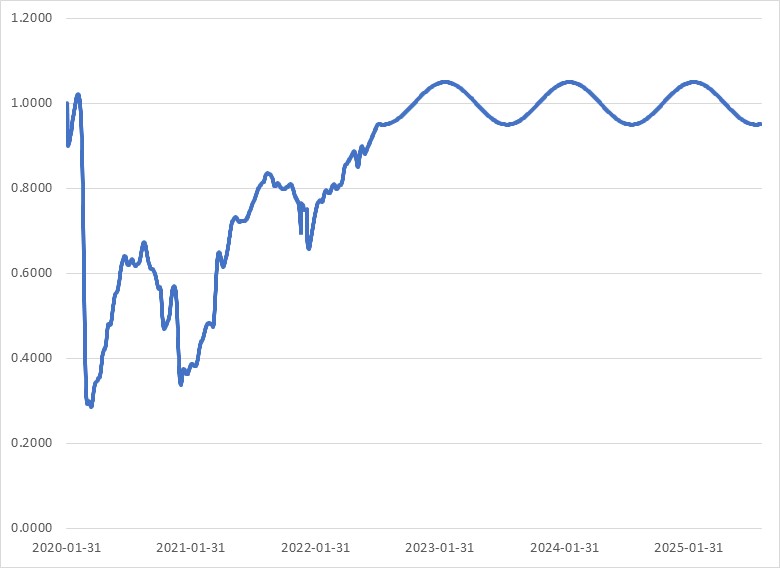

### 1.3. Reduction in effective contacts due to vaccination

#### 1.3.1. Overview of past UK vaccine campaigns

The model structure captures the different UK vaccine campaigns. Most older children and adults in the UK were eligible for a primary series plus one reinforcing dose (booster) of the COVID-19 vaccines.[5] In addition, selected populations deemed at high risk of severe consequences due to COVID-19 infection were eligible for additional campaigns as summarized in Table 3.

Table 3. Overview of the targeted population for past UK booster campaigns

| **Booster campaign** | **Target Population** |
| --- | --- |
| Spring booster 2022 | - Adults ages ≥75 years - Residents in care home for older adults - Individuals ages ≥12 years who are immunosuppressed |
| Autumn booster campaign 2022 | - Adults ages ≥50 years - Persons aged 5 – 49 years in a clinical risk group - Residents in care home for older adults - Staff working in care homes for older adults - Frontline health and social care workers - Persons aged 5 – 49 years who are household contacts of those with immune suppression - Persons aged 16 – 49 years who are carers |
| Spring 2023 | - Adults ages ≥75 years - Residents in care home for older adults - Individuals ages ≥5 years who are immunosuppressed |
| Autumn 2023 | - Adults ages ≥65 years - Persons aged 6 months – 64 years in a clinical risk group - Residents in care home for older adults - Staff working in care homes for older adults - Frontline health and social care workers - Persons aged 12 – 64 years who are household contacts of those with immune suppression - Persons aged 16 – 64 years who are carers |
| Spring 2024 | - Adults ages ≥75 years - Residents in care home for older adults - Individuals ages ≥ 6 months who are immunosuppressed |

#### 1.3.2. Vaccine coverage

Vaccine uptake was based on the weekly uptake data available for England as illustrated in Figure 2 through Figure 6.[6]

Figure 2. Percentage of the population who have completed their primary series, by age group

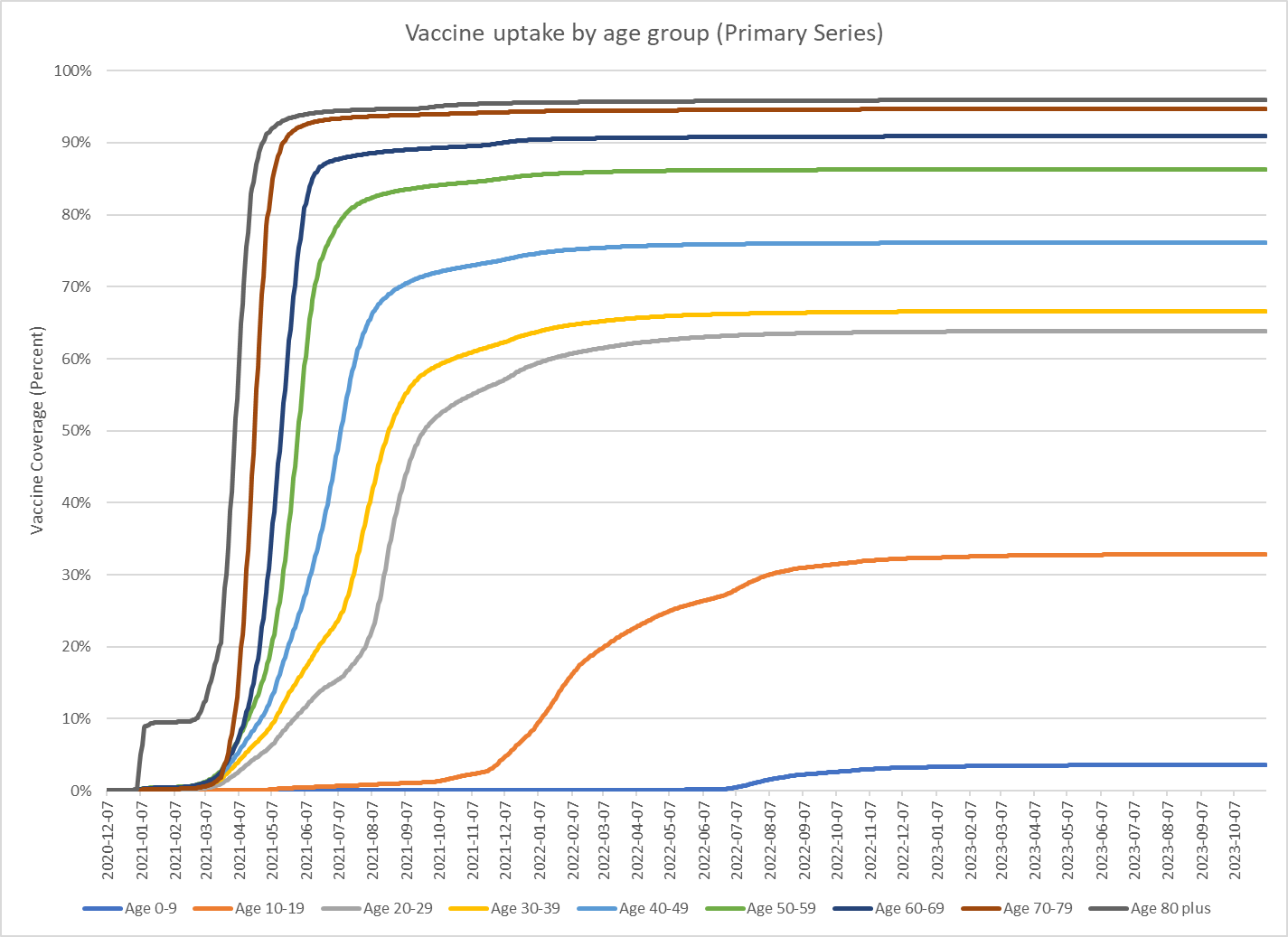

**Figure 3. Percentage of the population who have received the first booster, by age group**

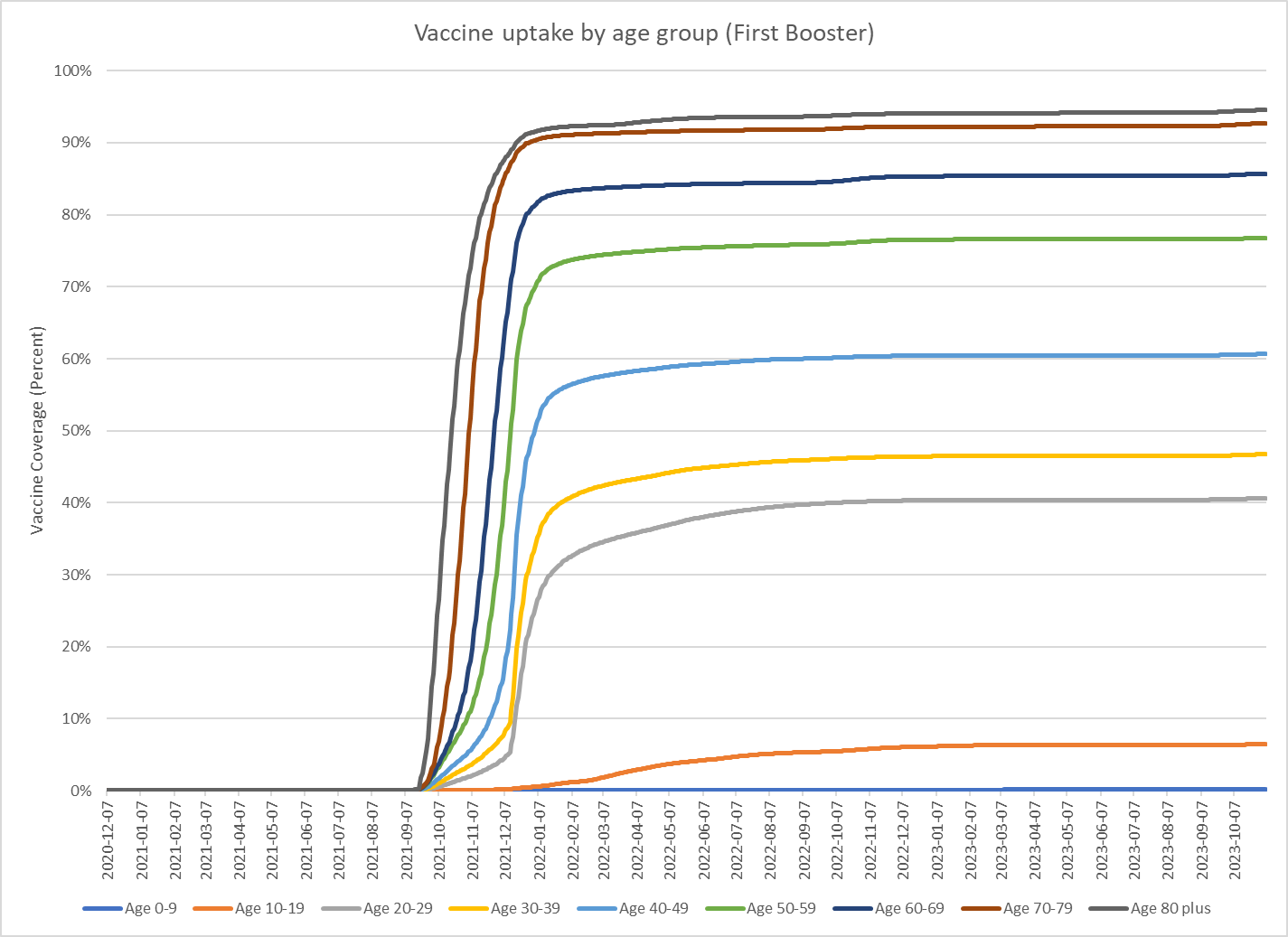

Figure 4. Percentage of the population who received the Spring 2022 booster, by age group

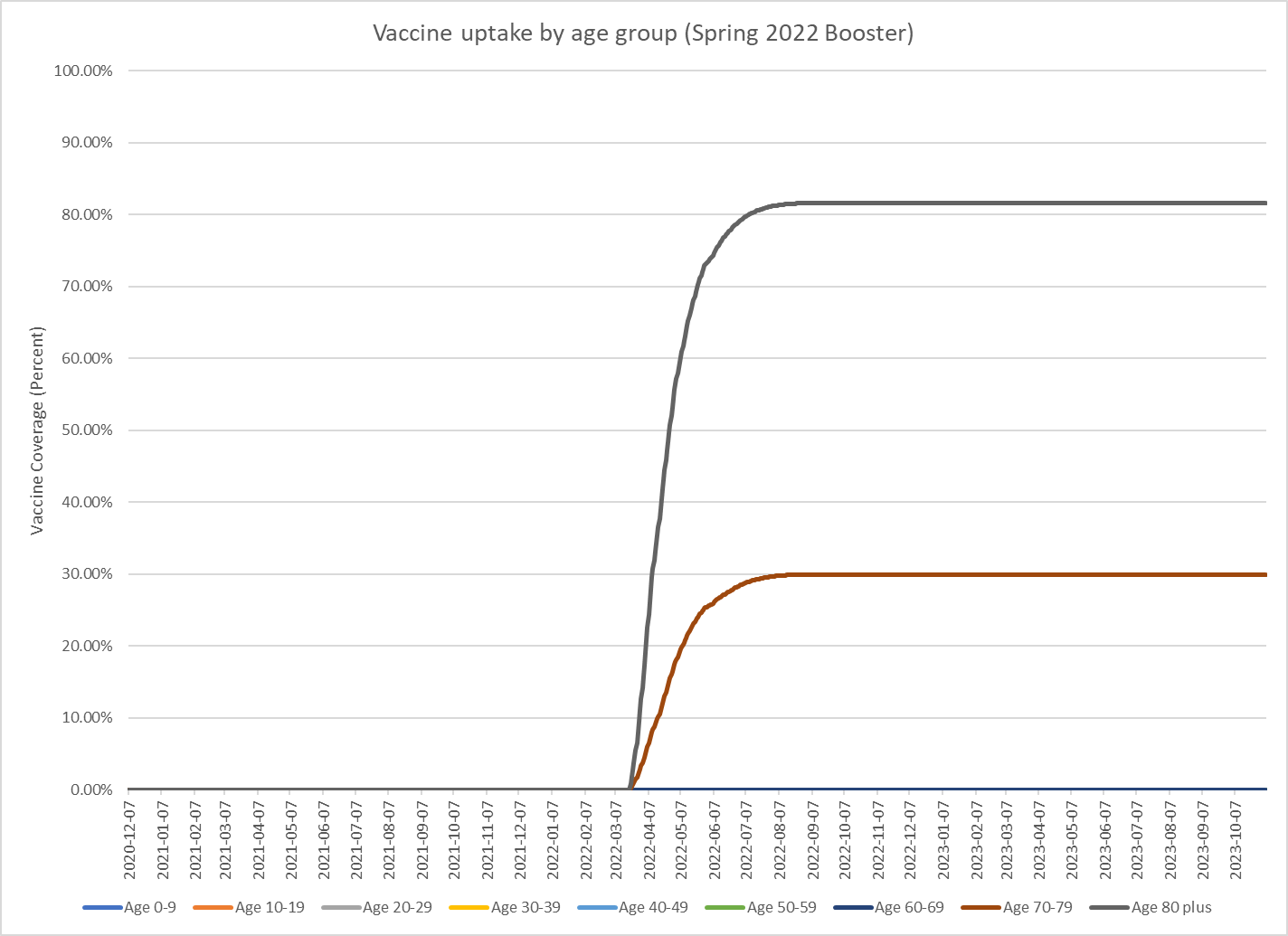

Figure 5. Percentage of the population who received the Autumn 2022 booster, by age group

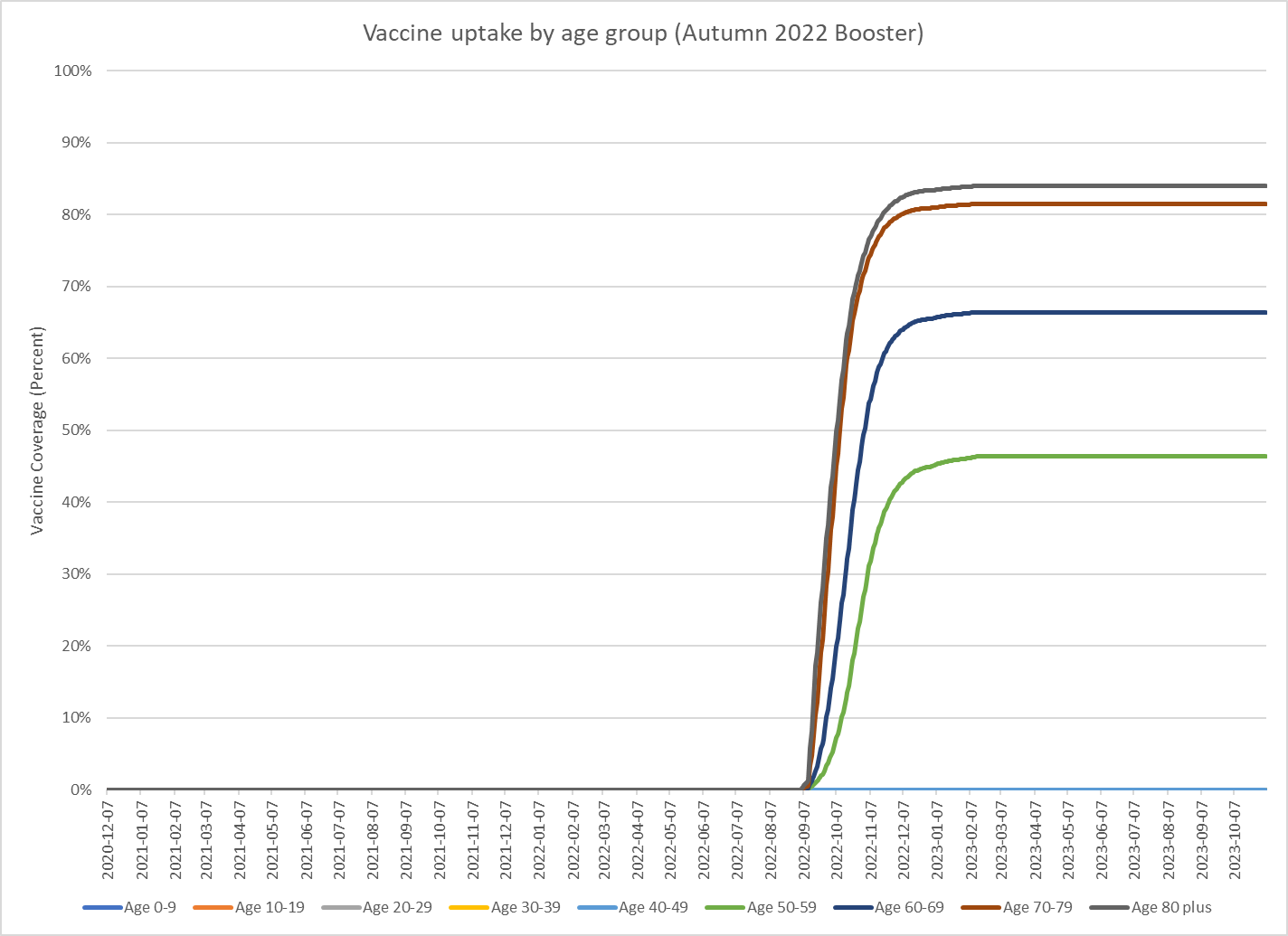

Figure 6. Percentage of the population who received the Spring 2023 booster, by age group

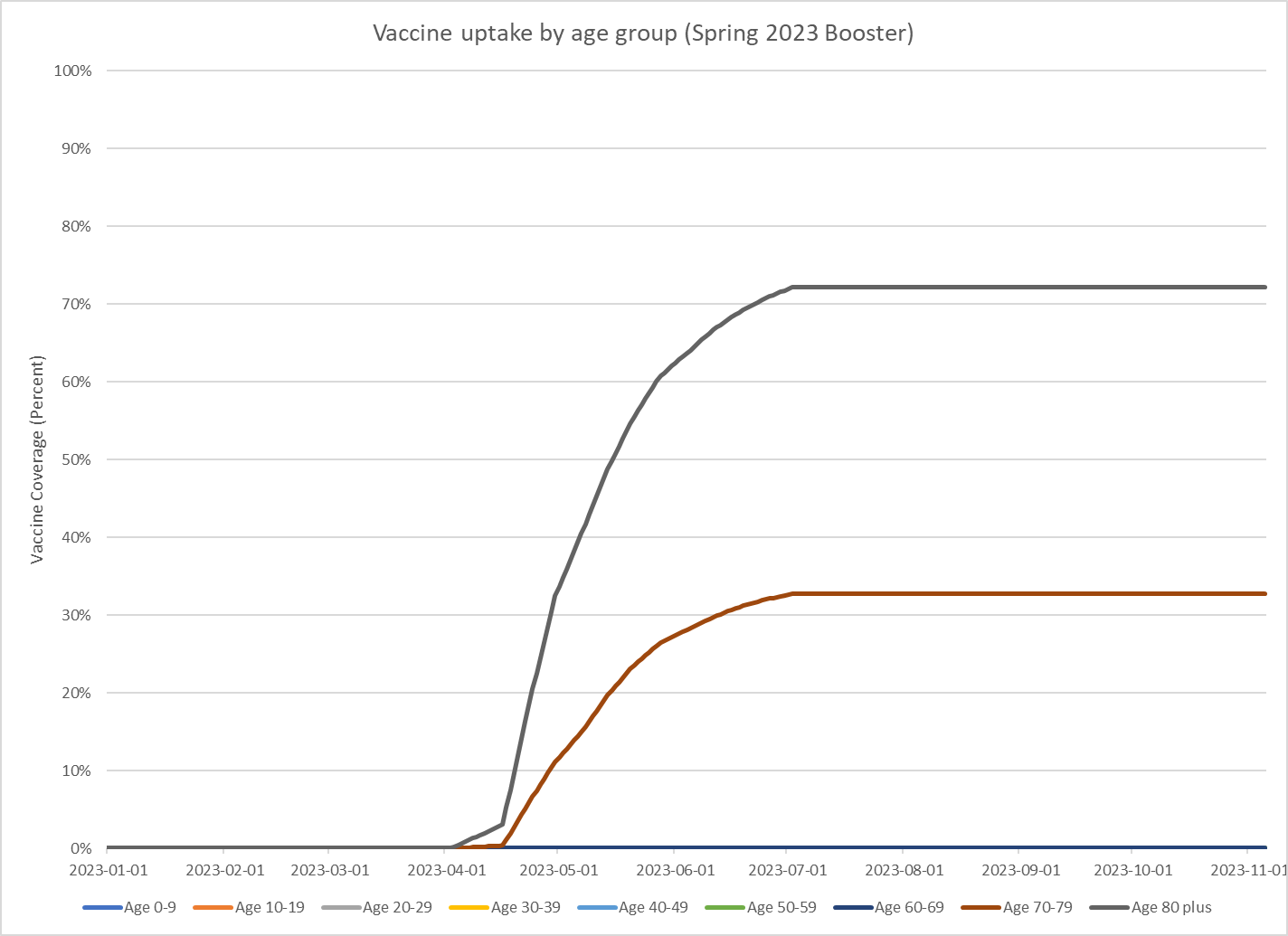

#### 1.3.3. Vaccine effectiveness

Initial vaccine effectiveness (VE) and monthly waning against infection and severe disease was required to determine the residual protection existing in the population due to previously received vaccines. These values differed by the vaccine, vaccination type (primary series or booster), and the circulating variant at the time of administration. Vaccine-specific VE and waning values are displayed in Table 4

Delta

Primary series and booster VEs during the Delta period for Spikevax and Comirnaty are described in the previously.[2] VEs for Vaxzevria (ChAdOX, Astra Zeneca) were not available, they were approximated by adjusting the primary series VE using the same ratio between primary series and booster observed with Comirnaty. Booster waning for infection and severe disease were assumed to be the same as primary series waning for infection and severe disease, respectively.

BA.1/BA.2

Primary series and booster VEs against BA.1 and BA.2 for Spikevax and Comirnaty, as well as waning for all boosters are described previously.[2] The VE against hospitalization for the Vaxzevria booster was obtained from the UKHA COVID-19 surveillance report.[7] Waning against primary series VE was obtained from Pratama et al. (2022).[8]

BA.4/BA.5

Booster VE against infection and hospitalization for monovalent and bivalent Spikevax and Comirnaty vaccines, as well as monthly waning, were estimated as described previously.[2]

XBB.1.5

For the XBB.1.5 boosters (Autumn 2023 and Spring 2024), VE was assumed to be the same as the Moderna Spikevax bivalent vaccine.

Table 4. Starting vaccine effectiveness and monthly waning values for previously-administered vaccinations

|  | **Primary series (%)** | | | | **Booster (%)** | | | |
| --- | --- | --- | --- | --- | --- | --- | --- | --- |
|  | **Infection** | | **Severe** | | **Infection** | | **Severe** | |
|  | **VE** | **Waning** | **VE** | **Waning** | **VE** | **Waning** | **VE** | **Waning** |
| **DELTA** |  |  |  |  |  |  |  |  |
| Spikevax | 91.0 | 3.4 | 97.0 | 1.0 | 93.3 | 3.4 | 95.7 | 1.0 |
| Comirnaty | 84.0 | 4.0 | 95.0 | 1.3 | 88.1 | 4.0 | 93.0 | 1.3 |
| Vaxzevria | 69.0 | 4.4 | 94.0 | 4.1 | 72.4 | 4.4 | 72.0 | 4.1 |
| **BA.1/BA.2** |  |  |  |  |  |  |  |  |
| Spikevax | 47.8 | 3.0 | 66.9 | 1.4 | 57.1 | 4.8 | 95.9 | 1.4 |
| Comirnaty | 52.2 | 3.4 | 80.2 | 1.4 | 53.0 | 4.8 | 89.1 | 1.4 |
| Vaxzevria | 56.0 | 8.5 | 85.0 | 5.6 | 61.1 | 4.8 | 90.0 | 1.4 |
| **BA.4/BA.5** |  |  |  |  |  |  |  |  |
| Spikevax |  |  |  |  | 32.3 | 4.8 | 62.6 | 1.4 |
| Comirnaty |  |  |  |  | 23.9 | 4.8 | 52.7 | 1.4 |
| Vaxzevria |  |  |  |  | 31.1 | 4.8 | 56.0 | 1.4 |
| Spikevax bivalent |  |  |  |  | 57.1 | 4.8 | 84.3 | 1.4 |
| Comirnaty bivalent |  |  |  |  | 53.0 | 4.8 | 82.6 | 1.4 |

VE, vaccine effectiveness.

Summary

Market share data for primary series and boosters were obtained from the Medicines & Healthcare products Regulatory Agency (MHRA) and used to calculate weighted VEs against infection and hospitalization.[9] Initial VEs and waning rates, weighted by market shares of each vaccine, which are entered into the SEIR model, are displayed in Table 5. Once again, for the XBB.1.5 boosters (Autumn 2023 and Spring 2024), VE was assumed to be the same as the Moderna Spikevax bivalent vaccine.

Table 5. Starting weighted vaccine effectiveness and monthly waning values for previously-administered vaccinations

|  | **Delta** | | | | **BA.1/BA.2** | | | | **BA.4/BA.5** | | | |
| --- | --- | --- | --- | --- | --- | --- | --- | --- | --- | --- | --- | --- |
|  | **Infection** | | **Severe** | | **Infection** | | **Severe** | | **Infection** | | **Severe** | |
|  | **VE** | **Waning** | **VE** | **Waning** | **VE** | **Waning** | **VE** | **Waning** | **VE** | **Waning** | **VE** | **Waning** |
| Primary series | 77.2 | 4.2 | 94.6 | 2.6 | 53.8 | 5.8 | 82.0 | 3.4 |  |  |  |  |
| First booster | 89.6 | 3.8 | 93.8 | 1.2 | 54.2 | 4.8 | 91.0 | 1.4 |  |  |  |  |
| Second booster (Spring 2022) |  |  |  |  |  |  |  |  | 26.3 | 4.8 | 55.6 | 1.4 |
| Third booster (Autumn 2022) |  |  |  |  |  |  |  |  | 54.9 | 4.8 | 83.4 | 1.4 |
| Fourth booster (Spring 2023) |  |  |  |  |  |  |  |  | 54.9 | 4.8 | 83.4 | 1.4 |

VE, vaccine effectiveness.

Impact of new variant on VE

In order to simulate the emergence a new variant, it was assumed that protection from both vaccine-mediated and natural immunity dropped immediately on the day that the new variant was considered dominant. The dates and the relative declines in immunity are summarized in Table 6. The drop in natural immunity was implemented as a proportion of individuals moving out of the R state back into the S state.

Table 6. Dates and decline in protection from vaccine and natural immunity due to variant change over time

|  | **Start of Omicron period** | **Start in Omicron BA.4/BA.5** | **Start in Omicron XBB period** |
| --- | --- | --- | --- |
| Date | December 20, 2021 | June 15, 2022 | March 13, 2023 |
| Drop in VE (infection) | Primary series: 30.2%, Boosters 1 & 2: 39.5% | 51.5% | 10% |
| Drop in VE (hospitalization) | Primary series: 13.3%, Boosters 1 & 2: 2.9% | 38.9%1 | 10% |
| Proportion moving from the R to the S state | 30.2% | 51.5% | 10% |

VE, vaccine effectiveness.

### 1.4. Transmissibility: February 2020 to August 2024

An initial 103 infections were used to seed or start the pandemic during the first 30 days of the burn-in period. This number is equal to the number of cases estimated by the Institute for Health Metrics and Evaluation (IHME) over the first 30 days.[10] The transmissibility parameter, which was allowed to vary daily, was estimated through a model calibration process to ensure that the model reflects cases of COVID-19 experienced in the UK from February 2020 through January 2024. The analytical choices for the calibration process were made considering the recommendations of Vanni and colleagues.[11] The transmissibility parameter was allowed to vary on a daily basis.

An initial model calibration was performed in a qualitative way by visually comparing the daily incidence of COVID cases predicted by the model and the values obtained from IHME[12] and the Office for National Statistics (ONS)[13] (start of the pandemic to October 27, 2022). For the second time period of the calibration, until January 30, 2024, the monthly estimates of the number of hospitalizations from the model was to differ by no more than 10% from the target number of hospitalizations for each month reported with National Health Service (NHS) England data,[14] inflated to represent the entire UK. The monthly calculated hospitalization rates per 100,000 population are displayed in Figure 7.

Figure 7. Monthly hospitalization rates for COVID-19 per 100,000 population, overall and by age

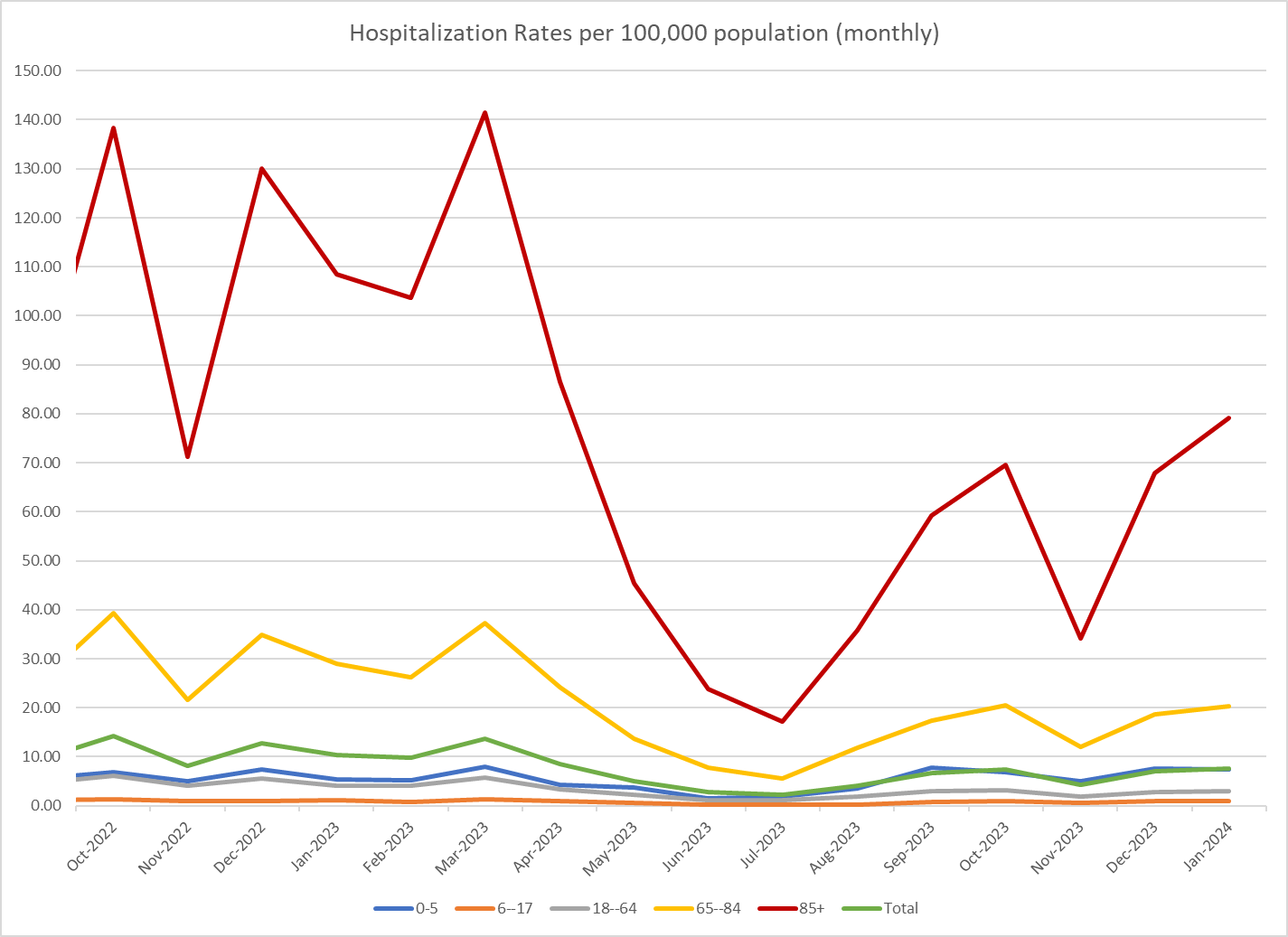

The final transmissibility parameters for the calibration period and the analytic time horizon for the base case scenario are summarized in Table 7. From January 2020 to January 2024, the transmissibility parameter was manually varied and linear interpolation was used to estimate values for the days between these dates. From February 2024 onwards assumptions were made as described in Section 2.2.

The comparison plot of the number of incident cases of COVID-19 predicted from the calibrated model and the corresponding estimates from the ONS and IHME is presented in Figure 8. The figures show that number of infections predicted by the model match the number of infections reported in the ONS and IHME period. A comparison of the number of hospitalizations actually reported and the number predicted by the model during the calibration period is presented in Table 8.

Table 7. Final transmissibility parameter inputs for the SEIR model

| **Date** | **Day from start of analysis** | **Transmissibility parameter estimate** |
| --- | --- | --- |
| January 31, 2020 | 1 | 0.20 |
| February 29, 2020 | 30 | 0.20 |
| March 15, 2020 | 45 | 0.40 |
| March 23, 2020 | 53 | 0.65 |
| March 31, 2020 | 61 | 0.65 |
| April 7, 2020 | 68 | 0.45 |
| April 15, 2020 | 76 | 0.29 |
| April 30, 2020 | 91 | 0.17 |
| May 31, 2020 | 122 | 0.11 |
| June 30, 2020 | 152 | 0.13 |
| July 31, 2020 | 183 | 0.15 |
| August 31, 2020 | 214 | 0.19 |
| September 30, 2020 | 244 | 0.23 |
| October 16, 2020 | 260 | 0.23 |
| October 31, 2020 | 275 | 0.16 |
| November 15, 2020 | 290 | 0.17 |
| November 30, 2020 | 305 | 0.13 |
| December 15, 2020 | 320 | 0.32 |
| December 31, 2020 | 336 | 0.26 |
| January 31, 2021 | 367 | 0.17 |
| February 28, 2021 | 395 | 0.14 |
| March 31, 2021 | 426 | 0.19 |
| April 30, 2021 | 456 | 0.12 |
| May 31, 2021 | 487 | 0.20 |
| June 15, 2021 | 502 | 0.29 |
| June 30, 2021 | 517 | 0.29 |
| July 16, 2021 | 533 | 0.19 |
| July 31, 2021 | 548 | 0.12 |
| August 15, 2021 | 563 | 0.18 |
| August 23, 2021 | 571 | 0.17 |
| August 31, 2021 | 579 | 0.15 |
| September 15, 2021 | 594 | 0.17 |
| September 21, 2021 | 600 | 0.20 |
| September 30, 2021 | 609 | 0.21 |
| October 6, 2021 | 615 | 0.24 |
| October 16, 2021 | 625 | 0.20 |
| October 31, 2021 | 640 | 0.13 |
| November 8, 2021 | 648 | 0.16 |
| November 15, 2021 | 655 | 0.20 |
| November 30, 2021 | 670 | 0.25 |
| December 7, 2021 | 677 | 0.25 |
| December 15, 2021 | 685 | 0.40 |
| December 23, 2021 | 693 | 0.35 |
| December 31, 2021 | 701 | 0.28 |
| January 9, 2022 | 710 | 0.15 |
| January 23, 2022 | 724 | 0.28 |
| January 31, 2022 | 732 | 0.25 |
| February 7, 2022 | 739 | 0.25 |
| February 15, 2022 | 747 | 0.22 |
| February 21, 2022 | 753 | 0.20 |
| February 28, 2022 | 760 | 0.32 |
| March 15, 2022 | 775 | 0.41 |
| March 31, 2022 | 791 | 0.37 |
| April 7, 2022 | 798 | 0.30 |
| April 15, 2022 | 806 | 0.30 |
| April 30, 2022 | 821 | 0.24 |
| May 15, 2022 | 836 | 0.24 |
| May 23, 2022 | 844 | 0.35 |
| May 31, 2022 | 852 | 0.45 |
| June 15, 2022 | 867 | 0.45 |
| June 16, 2022 | 868 | 0.40 |
| June 17, 2022 | 869 | 0.40 |
| June 22, 2022 | 874 | 0.29 |
| June 30, 2022 | 882 | 0.22 |
| July 14, 2022 | 896 | 0.17 |
| July 31, 2022 | 913 | 0.15 |
| August 15, 2022 | 928 | 0.16 |
| August 23, 2022 | 936 | 0.17 |
| August 31, 2022 | 944 | 0.22 |
| September 6, 2022 | 950 | 0.29 |
| September 15, 2022 | 959 | 0.30 |
| September 30, 2022 | 974 | 0.25 |
| October 15, 2022 | 989 | 0.22 |
| October 31, 2022 | 1005 | 0.20 |
| November 30, 2022 | 1035 | 0.33 |
| December 24, 2022 | 1059 | 0.42 |
| January 7, 2023 | 1073 | 0.25 |
| January 31, 2023 | 1097 | 0.25 |
| February 15, 2023 | 1112 | 0.46 |
| February 28, 2023 | 1125 | 0.35 |
| March 14, 2023 | 1139 | 0.37 |
| March 31, 2023 | 1156 | 0.22 |
| April 30, 2023 | 1186 | 0.26 |
| May 16, 2023 | 1202 | 0.26 |
| May 31, 2023 | 1217 | 0.18 |
| June 15, 2023 | 1232 | 0.30 |
| June 30, 2023 | 1247 | 0.29 |
| July 15, 2023 | 1262 | 0.20 |
| July 31, 2023 | 1278 | 0.17 |
| August 7, 2023 | 1285 | 0.44 |
| August 31, 2023 | 1309 | 0.33 |
| September 15, 2023 | 1324 | 0.23 |
| September 30, 2023 | 1339 | 0.23 |
| October 15, 2023 | 1354 | 0.35 |
| October 31, 2023 | 1370 | 0.30 |
| November 7, 2023 | 1377 | 0.16 |
| November 30, 2023 | 1400 | 0.18 |
| December 7, 2023 | 1407 | 0.40 |
| December 31, 2023 | 1431 | 0.33 |
| January 15, 2024 | 1446 | 0.20 |
| January 31, 2024 | 1462 | 0.20 |
| February 15, 2024 | 1477 | 0.46 |
| February 29, 2024 | 1491 | 0.35 |
| March 14, 2024 | 1505 | 0.37 |
| March 31, 2024 | 1522 | 0.22 |
| April 30, 2024 | 1552 | 0.26 |
| May 16, 2024 | 1568 | 0.26 |
| May 31, 2024 | 1583 | 0.18 |
| June 15, 2024 | 1598 | 0.30 |
| June 30, 2024 | 1613 | 0.29 |
| July 15, 2024 | 1628 | 0.20 |
| July 31, 2024 | 1644 | 0.17 |
| August 7, 2024 | 1651 | 0.44 |
| August 31, 2024 | 1675 | 0.33 |
| September 15, 2024 | 1690 | 0.23 |
| September 30, 2024 | 1705 | 0.23 |
| October 15, 2024 | 1720 | 0.35 |
| October 31, 2024 | 1736 | 0.30 |
| November 7, 2024 | 1743 | 0.16 |
| November 30, 2024 | 1766 | 0.18 |
| December 7, 2024 | 1773 | 0.40 |
| December 31, 2024 | 1797 | 0.33 |
| January 15, 2025 | 1812 | 0.20 |
| January 31, 2025 | 1828 | 0.20 |
| February 28, 2025 | 1856 | 0.20 |
| March 31, 2025 | 1887 | 0.20 |
| April 30, 2025 | 1917 | 0.20 |
| May 31, 2025 | 1948 | 0.20 |
| June 30, 2025 | 1978 | 0.20 |
| July 31, 2025 | 2009 | 0.20 |
| August 31, 2025 | 2040 | 0.20 |

Figure 8. Comparison of incidence of COVID-19 infections over time (calibrated model estimates versus ONS and IHME estimate) with the Moderna Autumn 2024 campaign

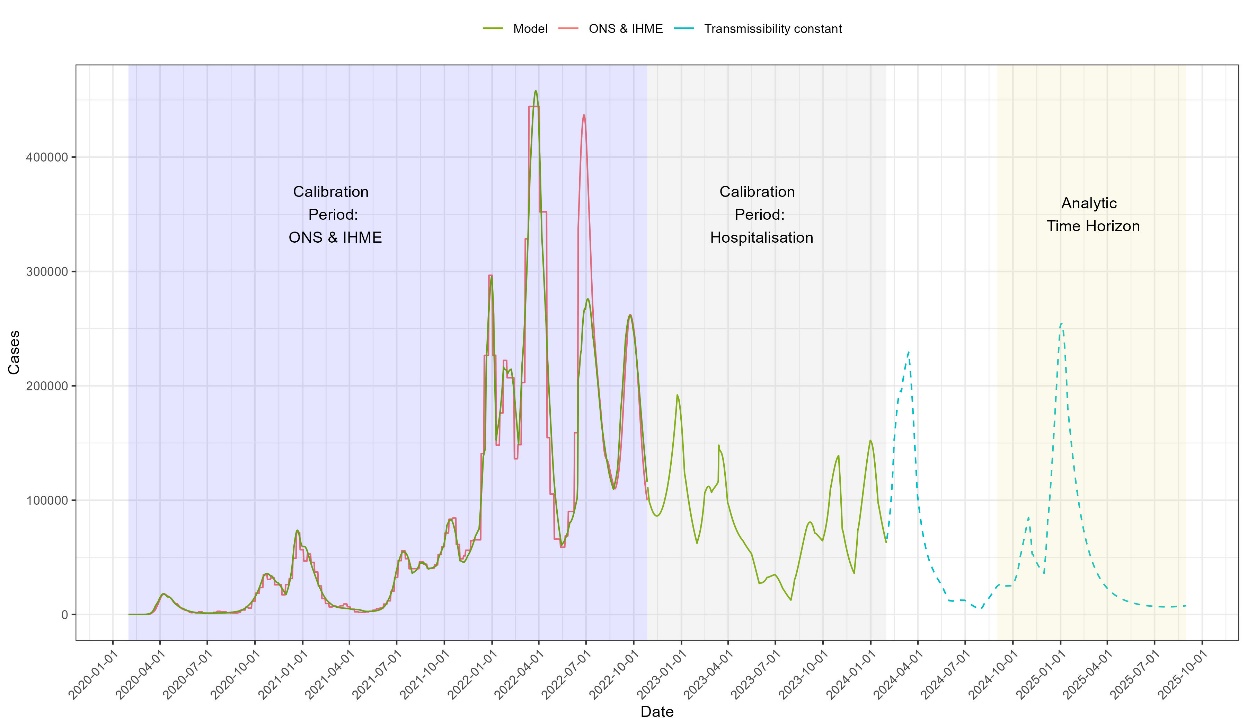

IHME, Institute for Health and Metrics and Evaluation; ONS, Office for National Statistics.

Table 8. Comparison of number of hospitalizations during the model burn-in period (model predicted versus observed from two past periods

| **Description** | | **No Autumn 2024 Campaign (all)** | **Moderna Autumn 2024 Campaign (all)** | **No Autumn 2024 Campaign (65+)** | **Moderna Autumn 2024 Campaign (65+)** |
| --- | --- | --- | --- | --- | --- |
| Target | Observed September 2022 to August 2023 | 67,662 | | 46,348 | |
| Alternative target | Observed February 2023 to January 2024 | 53,047 | | 36,039 | |

2. SEIR model inputs: analytic time period (September 2024 to August 2025)

The inputs that were changed for the analytic time period are described below. Inputs not described were the same as during the burn-in period.

### 2.1. Vaccine characteristics

The vaccine effectiveness and coverage were updated for the projection period and this has been described in the main manuscript. Figure 9 shows the pattern of uptake assumed for the Autumn 2024 campaign based on the Autumn 2023 campaign. Table 9 shows the final uptake values by age group.

Figure 9. COVID-19 vaccine uptake for the Autumn 2023 campaign in England, by age group

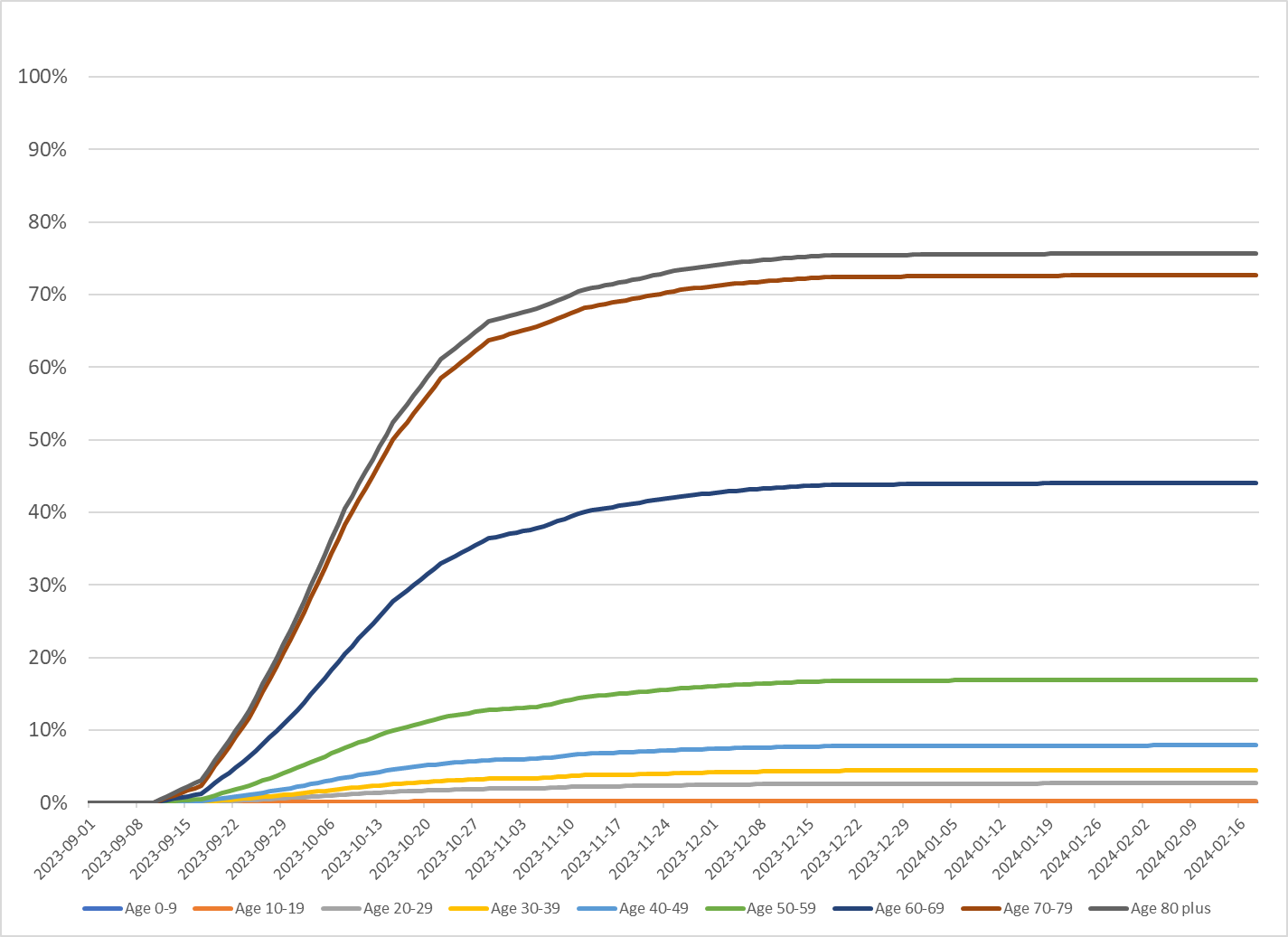

Table 9. Overall vaccine coverage rates assumed for the base case analysis

| **Age group (years)** | **Percent of age group who received the vaccine**[6] |
| --- | --- |
| 10 - 19 | 0.27% |
| 20 – 29 | 2.67% |
| 30 – 39 | 4.49% |
| 40 – 49 | 7.88% |
| 50 – 59 | 16.96% |
| 60 – 69 | 44.09% |
| 70 - 79 | 72.69% |
| 80+ | 75.71% |

### 2.2. Transmissibility parameter

As shown in Figure 10 for the base case scenario, the pattern from February 2023 to January 2024 was replicated for February 2024 to January 2025, and then held constant. The values for these projections, which cross the burn-in and the analytic time horizon, were displayed in Table 7.

Figure 10. Transmissibility across the simulation time horizon: values for the base case

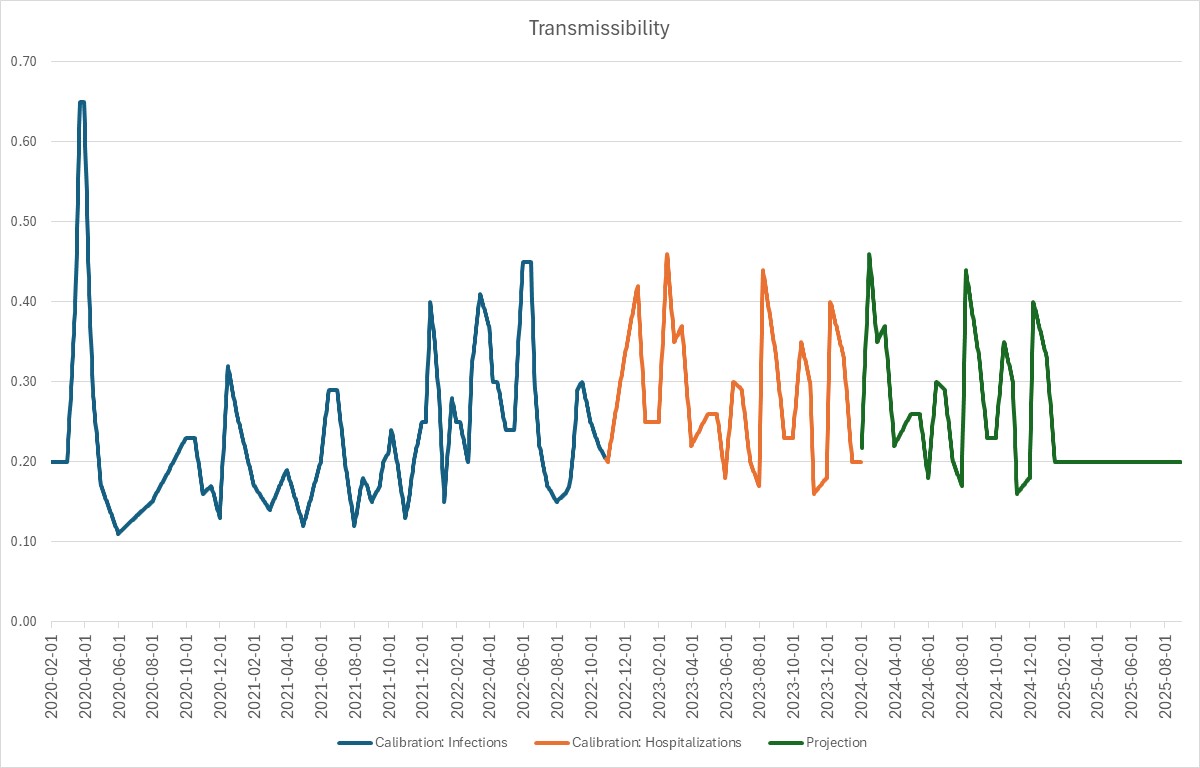

Figure 11. Transmissibility across the simulation time horizon: values for the alternative scenario

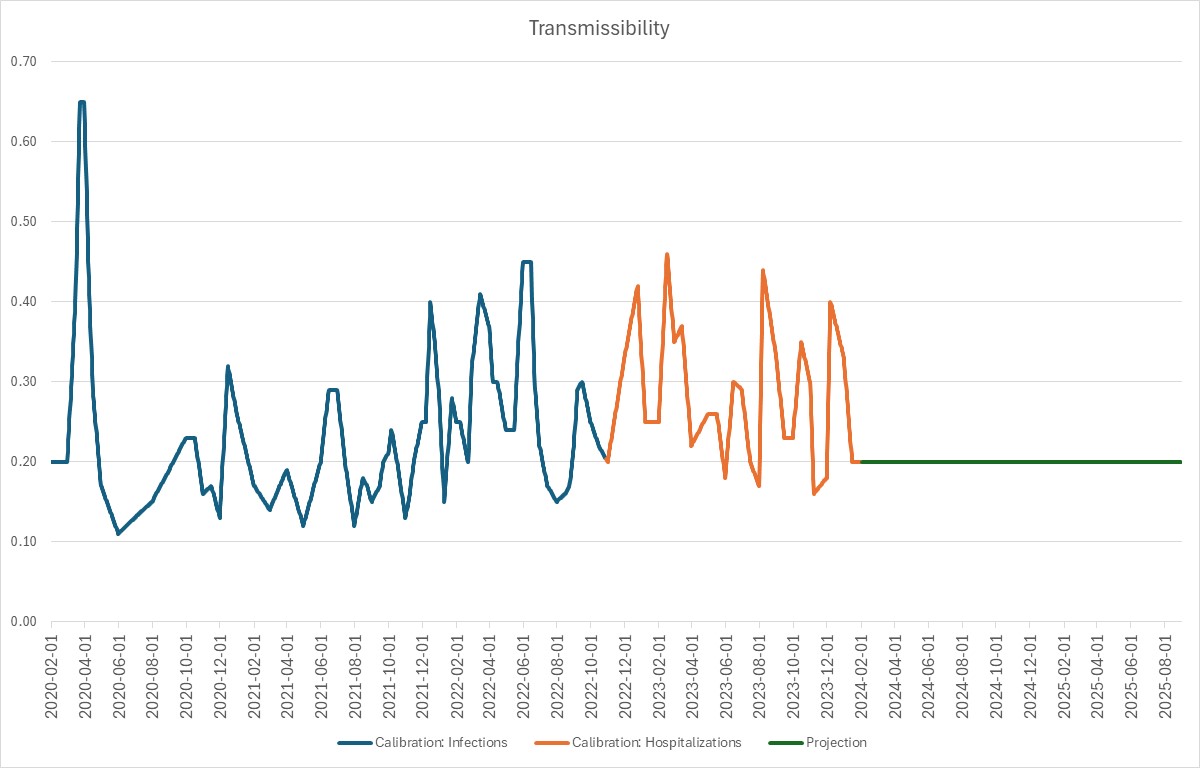

3. Infection consequences model

Additional information on the inputs for the infection consequences model are provided in this section. A schematic describing the decision tree is provided in Figure 12.

As mentioned in the main manuscript, the number of SARS-CoV-2 infections (both asymptomatic and symptomatic) are predicted by the SEIR model, and entered into the infection consequences model. For all symptomatic infections, there is a probability, dependent on age, of requiring hospitalization. As the SEIR model does not track infections by risk of severe outcomes, the consequences model also cannot stratify individuals by risk of severe disease and therefore the probability of hospitalization only varies by age. Vaccination reduces this probability, with the VE calculated within the SEIR model and varying by strata. Those who are hospitalized face a risk of mortality.[15] There is also an increased risk of mortality post-discharge compared to general population age-specific mortality rates,[16] as well as a risk of hospital readmission.[17] Those that survive hospitalization receive a toll, to account for additional health care resource use and QoL decrements following a SARS-CoV-2 infection. Survivors are also at risk for long COVID,[18] and can incur costs and QALY decrements.

Symptomatic SARS-CoV-2 infected patients not requiring hospitalization are divided into two groups: i) those who seek outpatient medical care (include outpatient physician visits or emergency room visits without hospital admissions);[19] and ii) those who do not seek medical care. The decision tree structure does not differentiate between these two groups, however, the inputs and calculations for the associated health care costs are structured to allow for this difference. Cost inputs for outpatient care are weighted by the proportion of patients who seek medical care, with a cost of zero assigned to those who do not seek care. Similar to inpatients, a post-infection toll is applied. Non-hospitalized patients are considered to have a non-severe infection and are not at risk of death from COVID-19, however, a risk for long COVID is applied.

Also not shown in the decision tree is that there is a risk for infection-related myocarditis for all infected individuals.[20] This is applied as a “toll” to capture the additional costs and short-term QoL impact associated with the small percentage of individuals who develop this complication. This event is independent from future events and does not impact the probability of other related events in the decision tree. This is a conservative approach as the Autumn 2024 Campaign is expected to reduce the number of infections compared to a No Autumn 2024 Campaign and, therefore, reduce the number of cases of infection-related myocarditis.

Figure 12. Illustration of the infection consequences decision tree

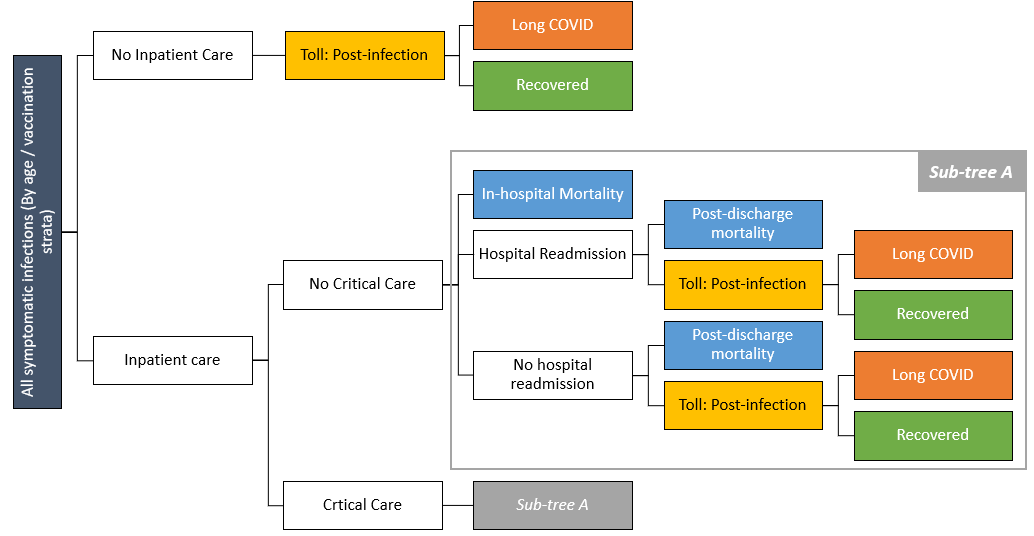

### 3.1. Percent with symptomatic disease

For the base case, the proportion of infections that are asymptomatic (25%) was obtained from a global systematic review and meta-analysis by Alene et al.[21] As values were not disaggregated by age, data on the number of COVID-19 hospitalizations from January 2022 to October 2022[14] and the model output on number of infections were used to estimate age-specific proportions. The number of infections had been previously calibrated to predict the number of total infections from January 2022 to October 2022. When it was assumed that 75% of infections across all age groups were symptomatic, the model predicted too many hospitalizations for this period in the younger age groups and too few for older adults. This input value was therefore adjusted by age group so that the model-predicted hospitalizations by age group approximated the observed totals (Table 10). The lower rate of symptomatic disease in younger populations may be due to detection bias; these age groups with mild symptoms may no longer test for COVID-19 and also may no longer visit healthcare providers when they do receive a positive test. Therefore, many milder infections in the younger age groups may not come to medical attention and are not counted in the denominator in studies that generate hospitalization rates.

Table 10. Developing age-specific symptom rates: observed and model-predicted hospitalizations, January 2022 to October 2022

| **Target data age group** | **Observed target^a^** | **Model-predicted** |
| --- | --- | --- |
| 0-5 years | 5,400 | 4,861 |
| 6-17 years | 3,015 | 3,189 |
| 18-64 years | 47,331 | 41,734 |
| 65-84 years | 51,757 | 45,258 |
| 85+ years | 25,830 | 15,593 |
| **Total** | **133,334** | **110,635** |

^a^Target = number of COVID-19 hospitalizations, by age group, reported for the period of January 2022 to October 2022.[14]

### 3.2. Long COVID quality-adjusted life-year (QALY) loss

In the base case, the QALY toll associated with long COVID was set to 0 in order to be conservative and avoid double-counting with the post-infection QALY toll. As the post-infection toll accounted for only 48 days for hospitalized patients and 6 months for non-hospitalized patients, it does account for the full loss of QALYs for those with long COVID. Therefore a sensitivity analysis was conducted using the long COVID QALY toll of 0.259 for hospitalized patients and 0.133[22] for non-hospitalized patients. These values were based on NICE EAG's preferred value in TA900 based on COVID-19 data after 5 months (Evans et al.[23]) and adjusted to cover 1 year using results from Evans et al.[24] It was assumed those not hospitalized had World Health Organization (WHO) severity 3-5 (i.e., not hospitalized/low flow oxygen). A weighted average was calculated using patient numbers reported by Evans et al.[25] in the "unsure" or "not recovered" COVID-19 groups.

### 3.3. Opportunity costs

In the Department of Health report on the impact of Autumn 2023 boosters,[26] the effect of COVID-19 hospitalizations on elective procedures was estimated because the NHS is still in a “recovery phase” from COVID-19. Hence, in our analysis, the additional benefit of reducing COVID-19 hospitalizations was assessed by calculating the opportunity cost of bed days required for other hospitalizations using a similar approach to Brassel et al. and Sandmann et al.[27,28] The opportunity cost per COVID-19 hospitalization is the sum of the hospital cost for COVID-19 and the net monetary benefit forgone from the elective hospitalization. The formula used to calculate the opportunity cost is presented below.

$$Opportunity cost= {Cost}_{COVID19}+\left( \frac{{LOS}_{COVID19}}{{LOS}_{Elective}} \right)\times\left( {Benefit}_{Elective}*WTP-{Cost}_{Elective} \right)$$

Where:

${LOS}_{COVID19}$: Length of hospital stay of a COVID-19 hospitalization

${LOS}_{Elective}$: Length of hospital stay of an elective, non-COVID-19 treatment

${Benefit}_{Elective}$: Benefit (in QALYs) for an elective, non-COVID-19 treatment

$WTP$: Willingness to pay

${Cost}_{COVID19}$: Average cost of COVID-19 hospitalization

${Cost}_{Elective}$: Average cost of elective care treatment

Inclusion of opportunity costs in the base case analysis assumes that hospital beds can be made available to treat other patients not hospitalized with COVID-19 infection. Other aspects of healthcare system strengthening and reducing crowded hospitals especially during the winter months has not been considered in the analysis.

### 3.4. Societal perspective

Table 11 includes the additional inputs for the societal perspective and key assumptions utilized in this scenario analysis, including time loss from work due to acute infection and hospitalization recovery. The proportions of patients with long COVID and severe long COVID are also presented, as severe long COVID patients are assumed to experience time loss.

Table 11. Additional (non–healthcare-related) inputs for the societal perspective scenario analysis

| **Model parameter** | **Value** |
| --- | --- |
| Percentage in labor force[29] |  |
| 18-29 years | 70.37% |
| 30-39 years | 85.41% |
| 40-49 years | 85.24% |
| 50-64 years | 71.02% |
| 65-74 years | 11.53% |
| 75-84 years | 11.53% |
| ≥85 years | 11.53% |
| Daily wage[30,31] | £131.90 |
| Days lost^a^ |  |
| Vaccination^b^[32,33] | 0.04 |
| Symptomatic infection, not hospitalized^c^[34] | 3.57 |
| Symptomatic infection, hospitalized^d^ |  |
| No critical care[35] | 6.26 |
| Critical care[35] | 8.96 |
| Hospitalization recovery[36] | 13.90 |
| Any long COVID[37] | 4.46 |
| Severe long COVID[38] | 9.34 |
| Infection-related myocarditis[39] | 6.67 |
| Adverse events |  |
| Grade 3 local[40] | 0.75 |
| Grade 3 systemic[40] | 0.75 |
| Anaphylaxis[39] | 1.12 |
| Vaccine-induced myocarditis/pericarditis[39] | 3.66 |

^a^Days lost may not fall only on work days; estimates were adjusted by 5/7 to account for this.

^b^Assumes 0.25 hours, assuming a 6.64-hour work day.

^c^Applies to all symptomatic non-hospitalized patients, including those who did not seek outpatient care. Assumes 5 days.

^d^Time loss due to symptomatic infection, not hospitalized, added to displayed values to account for symptomatic period prior to hospitalization.

### 3.5. Vaccine-related adverse events

It was assumed that there are no differences in the adverse event (AE) rates between the Moderna Autumn 2024 vaccine and prior versions. AEs rates were obtained from the Moderna mRNA-1273 clinical trial report[41] and represent solicited AEs that occurred within 7 days of receiving the 50 µg monovalent booster injection. Grade 0, 1, and 2 AEs (both local and systemic) are expected to be relatively minor in nature, and as such are unlikely to result in healthcare resource utilization. Accordingly, the model included grade 3 local and systemic AEs only In the trial, there were 5.6% of patients who experienced any grade 3 local AE and 12.9% who experienced any grade 3 systemic AE. Overall, it was reported that a total of 17.6% of patients experienced any type of Grade 3 AE, suggesting that 0.7% of patients may have experienced both a local and systemic AE. To avoid double-counting physician visits, the 5.6% of patients experiencing a grade 3 local AE was reduced by 0.7% to 4.9%, and this value was used in the model. Further, although it was reported that 12.9% of booster recipients experienced a grade 3 systemic AE, placebo patients also reported experiencing grade 3 systemic AEs. In the clinical trial comparing the primary series (dose 1 and 2) to placebo, on average, those receiving placebo experienced 41% fewer systemic AEs than those who received mRNA-1273. Therefore, to adjust for background systemic AEs that may be attributed to the placebo, the 12.9% was multiplied by 59%, resulting in a value of 7.6%. This value is used in the model for the estimation of the proportion of patients experiencing any grade 3 systemic AE.

The probability of anaphylaxis (per dose) from a mRNA vaccine is based on a chart review of cases individuals receiving primary series doses.[42] The number of cases/million doses was 4.9 and 5.0 for Moderna and Pfizer-BioNTech, respectively. Because there was no statistically significant difference between the vaccines, an average value of 5.0/million doses was used for both vaccines.

The rate of vaccine-induced myocarditis/pericarditis (per dose) in individuals age 18-39 years was obtained from an analysis of Vaccine Safety Datalink (VSD) data.[43] Cases of myocarditis and pericarditis during 0-7 days post first booster vaccination with Moderna and Pfizer-BioNTech monovalent were compared to cases during the comparison interval (22-42 days post first booster vaccination). The excess number of events/million doses administered was 30.9 for males, and 4.9 for females. Although separate values for Moderna and Pfizer-BioNTech were available, pooled values were used, as there were no statistically significant differences between the two mRNA vaccines. The average of these pooled values was calculated to obtain a value of 17.9 excess events/million doses administered (0.00179%). Model input parameters related to vaccine-related AEs are displayed in Table 12.

Table 12. Model inputs for the vaccine-related adverse events: base case values

| **Model parameter** | **Value** |
| --- | --- |
| Rates | |
| Grade 3 local^a^[41] | 4.90% |
| Grade 3 systemic^a^[41] | 7.61% |
| Anaphylaxis^b^[42] | 0.0005% |
| Vaccine-induced myocarditis/pericarditis[44] | 0.0018% |
| Costs | |
| Grade 3 local | £0.00 |
| Grade 3 systemic | £4.04 |
| Anaphylaxis | £439.22 |
| Vaccine-induced myocarditis/pericarditis[45,46] | £3,972.45 |
| QALYs lost[47] | |
| Grade 3 local | 0.0003 |
| Grade 3 systemic | 0.0011 |
| Anaphylaxis | 0.0019 |
| Vaccine-induced myocarditis/pericarditis | 0.0019 |

QALY, quality-adjusted life-year.

^a^Rates are from the AEs for monovalent boosters and assumed to apply to future vaccine versions. Grade 4 AEs were not included (there were no grade 4 AEs [local or systemic] reported in the clinical trial).

^b^Risk applies to those ages 18-39 years only.

4. Additional results tables and figures

### 4.1. Base case QALY and economic outcomes, disaggregated

Table 13. Base case: QALY and economic outcomes with and without the Moderna Autumn 2024 Campaign

| **Outcome** | **No Autumn 2024 Vaccine** | **Moderna Autumn 2024 Vaccine** | **Difference** | **Percent change** |
| --- | --- | --- | --- | --- |
| **QALYs lost** | | | | |
| QALYs lost - death | 113,852 | 81,320 | -32,532 | -29% |
| Not hospitalized | 35,264 | 28,562 | -6,702 | -19% |
| Hospitalized | 1,820 | 1,125 | -695 | -38% |
| Adverse events | 0 | 1,260 | 1,260 | -- |
| Infection-related myocarditis | 42 | 31 | -11 | -26% |
| Post-infection | 201,601 | 162,358 | -39,244 | -19% |
| QALYs lost - morbidity | 238,727 | 193,335 | -45,392 | -19% |
| *Total* | *352,580* | *274,655* | *-77,925* | *-22%* |
| **Costs (millions)** | | | | |
| Vaccination | £0 | £954 | £954 |  |
| Adverse events | £0 | £4 | £4 |  |
| Short-term infection costs | £479 | £318 | -£161 | -34% |
| Post-infection costs | £117 | £95 | -£22 | -19% |
| Infection-related myocarditis | £47 | £36 | -£11 | -24% |
| Long COVID | £182 | £147 | -£35 | -19% |
| Hospital opportunity costs | £163 | £101 | -£63 | -38% |
| *Total* | £989 | £1,654 | £665 | *67%* |

QALY, quality-adjusted life-year

### 4.2. Deterministic sensitivity analysis results

Table 14. DSA results (Moderna Autumn 2024 Campaign relative to No Autumn 2024 Campaign)

| **Variable** | **Base case value** | **Sensitivity analysis value** | **Incremental QALYs** | **Incremental cost** | **ICER** | **Impact on ICER** |
| --- | --- | --- | --- | --- | --- | --- |
| **Base Case** |  |  | **77,925** | **£665,447,770** | **£8,540** |  |
| Transmissibility | Transmissibility: same pattern as February 2023 to January 2024 for February 2024 to January 2025 | Transmissibility: constant for February 2024 onwards | 74,235 | £654,415,857 | £8,815 | 3% |
|  |  | Transmissibility +10% for February 2024 onwards | 81,302 | £648,438,086 | £7,976 | -7% |
|  |  | Transmissibility -10% for February 2024 onwards | 72,039 | £691,429,483 | £9,598 | 12% |
| Immune escape | No immune escape during analytic time period | Peak in June 2025 | 73,625 | £610,281,693 | £8,289 | -3% |
| Target population | Age 65+ | Age 50+ | 93,734 | £943,083,665 | £10,061 | 18% |
| Vaccine coverage |  | -10% | 68,893 | £601,215,005 | £8,727 | 2% |
| Moderna vaccine initial VE | Infection: 57.1% Hospitalization: 84.3% | Infection: 30.6% Hospitalization: 80.3% | 37,843 | £783,316,944 | £20,699 | 142% |
|  |  | Infection: 83.7% Hospitalization: 87.5% | 116,527 | £555,566,917 | £4,768 | -44% |
| Moderna vaccine initial VE: infection | Infection: 57.1% | Infection: 30.6% | 40,425 | £767,613,433 | £18,988 | 122% |
|  |  | Infection: 83.7% | 115,088 | £564,046,340 | £4,901 | -43% |
| Moderna vaccine initial VE: hospitalization | Hospitalization: 84.3% | Hospitalization: 80.3% | 75,738 | £678,546,144 | £8,959 | 5% |
|  |  | Hospitalization: 87.5% | 79,674 | £654,969,070 | £8,221 | -4% |
| Moderna vaccine VE: waning | Infection (monthly): 4.8% Hospitalization (monthly): 1.4% | Infection (monthly): 3.1% Hospitalization (monthly): 0.6% | 88,261 | £632,832,255 | £7,170 | -16% |
|  |  | Infection (monthly): 6.8% Hospitalization (monthly): 2.4% | 65,196 | £706,525,279 | £10,837 | 27% |
| Symptomatic infections | 0-4 years: 24.75% 5-17 years: 9.75% 18-29 years: 37.5% 30-39 years: 37.5% 40-49 years: 37.5% 50-64 years: 37.5% 65-74 years: 75% 75-84 years: 75% 85+ years: 93.75% | -10% | 70,006 | £694,686,393 | £9,923 | 16% |
|  |  | +10% | 85,843 | £636,209,146 | £7,411 | -13% |
| Hospitalization rates in the unvaccinated | 0-4 years: 0.51% 5-17 years: 0.35% 18-29 years: 0.52% 30-39 years: 0.47% 40-49 years: 0.28% 50-64 years: 0.48% 65-74 years: 1.79% 75-84 years: 6.02% 85+ years: 9.22% | Alternative Scenario (Shiri et al.[48]) | 90,782 | £618,863,872 | £6,817 | -20% |
|  |  | -25% | 69,332 | £715,607,232 | £10,321 | 21% |
|  |  | +25% | 86,517 | £615,288,307 | £7,112 | -17% |
| Hospital mortality rate | 0-4 years: 0.09% 5-17 years: 0.19% 18-29 years: 0.77% 30-39 years: 1.14% 40-49 years: 2.48% 50-64 years: 4.17% 65-74 years: 7.84% 75-84 years: 9.51% 85+ years: 17.04% | Alternative scenario (Shiri et al.[49]) | 93,069 | £668,551,936 | £7,183 | -16% |
|  |  | -25% | 75,013 | £664,771,413 | £8,862 | 4% |
|  |  | +25% | 80,836 | £666,124,126 | £8,240 | -4% |
| Hospital readmission rate | 13.5% | 10.1% | 77,901 | £668,966,716 | £8,587 | 1% |
|  |  | 16.9% | 77,948 | £661,928,823 | £8,492 | -1% |
| Post-discharge mortality | No critical care: 12.7% Critical care: 8.8% | -25% | 73,247 | £665,423,041 | £9,085 | 6% |
|  |  | +25% | 82,602 | £665,472,498 | £8,056 | -6% |
| Infection-related myocarditis | 0-4 years: 0.13% 5-17 years: 0.13% 18-29 years: 0.08% 30-39 years: 0.07% 40-49 years: 0.10% 50-64 years: 0.14% 65-74 years: 0.17% 75-84 years: 0.22% 85+ years: 0.22% | -25% | 77,922 | £668,312,526 | £8,577 | 0% |
|  |  | +25% | 77,927 | £662,583,013 | £8,503 | 0% |
| Rates of long COVID | 3.72% | -25% | 77,925 | £674,255,186 | £8,653 | 1% |
|  |  | +25% | 77,925 | £656,640,353 | £8,427 | -1% |
| Hospital costs | No critical care: £2,242.03 Critical care: £24,856.24 | -25% | 77,925 | £727,341,341 | £9,334 | 9% |
|  |  | +25% | 77,925 | £603,554,198 | £7,745 | -9% |
| Hospitalization recovery cost | £329.60 | -25% | 77,925 | £666,910,676 | £8,558 | 0% |
|  |  | +25% | 77,925 | £663,984,863 | £8,521 | 0% |
| Outpatient treatment cost | £40.49 | -25% | 77,925 | £666,041,859 | £8,547 | 0% |
|  |  | +25% | 77,925 | £664,853,680 | £8,532 | 0% |
| Long COVID costs | £67.68 | -25% | 77,925 | £666,439,177 | £8,552 | 0% |
|  |  | +25% | 77,925 | £664,456,363 | £8,527 | 0% |
| Adverse event costs | Grade 3 local: £0.00 Grade 3 systemic: £4.04 Anaphylaxis: £439.22 Myocarditis/pericarditis: £3,972.45 | -25% | 77,925 | £664,446,042 | £8,527 | 0% |
|  |  | +25% | 77,925 | £666,449,497 | £8,552 | 0% |
| Short-term infection period QALY loss (outpatient and hospitalizations) | Not hospitalized: 0.0043 Hospitalized, not critical care: 0.0168 Hospitalized, critical care: 0.033 | -25% | 76,075 | £665,447,770 | £8,747 | 2% |
|  |  | +25% | 79,774 | £665,447,770 | £8,342 | -2% |
| Adverse event QALY loss | Grade 3 local: 0.0003 Grade 3 systemic: 0.0011 Anaphylaxis: 0.0019 Myocarditis/pericarditis: 0.0019 | -25% | 78,239 | £665,447,770 | £8,505 | 0% |
|  |  | +25% | 77,610 | £665,447,770 | £8,574 | 0% |
| Post-infection QALY loss | Not hospitalized: 0.024 Hospitalized: 0.0911 | -25% | 68,114 | £665,447,770 | £9,770 | 14% |
|  |  | +25% | 87,735 | £665,447,770 | £7,585 | -11% |
| Vaccine-related myocarditis rates | 0.00177% | -25% | 77,925 | £665,436,613 | £8,539 | 0% |
|  |  | +25% | 77,925 | £665,458,926 | £8,540 | 0% |
| Perspective | Health care + opportunity loss due to bed days | Societal | 77,925 | £489,645,493 | £6,284 | -26% |
|  |  | Healthcare | 77,925 | £728,090,278 | £9,344 | 9% |
| % seeking care | 34.91% | -25% | 77,925 | £670,946,413 | £8,610 | 1% |
|  |  | +25% | 77,925 | £659,949,126 | £8,469 | -1% |
| Long COVID QALY | Excluded | Included | 85,815 | £665,447,770 | £7,754 | -9% |
| Discount rate | 3.5% | 0% | 89,143 | £665,447,770 | £7,465 | -13% |
| Comparator | No Autumn 2024 Vaccine | Pfizer-BioNTech (VE: meta-analysis[50]) | 29,184 | -£58,428,679 | Moderna Autumn 2024 Campaign dominates Pfizer-BioNTech-Autumn 2024 Campaign | n/a |

ICER, incremental cost-effectiveness ratio; n/a, not applicable; QALY, quality-adjusted life-year; VE, vaccine efficacy.

Figure 13. Number of daily infections over time for base case and four scenarios.

The number of daily infections projected to occur from September 1, 2024 to August 31, 2025 with each of the incidence scenarios is illustrated without and without the Autumn 2024 Campaign (See Technical Appendix Figure 13). The immune escape scenario has an extra incidence peak in June 2025 and the highest number of hospitalizations of all incidence scenarios (Table 15). The alternative scenario has a single wide peak, which means that when vaccination is implemented, there are still over 58,000 hospitalizations. This number is slightly higher than the 53,000 hospitalizations observed from February 2023 to January 2024. All other incidence scenarios predict less than 50,000 hospitalizations occurring from September 2024 to August 2023 with the Autumn 2024 vaccine campaign.

**A: No Autumn 2024 Campaign**

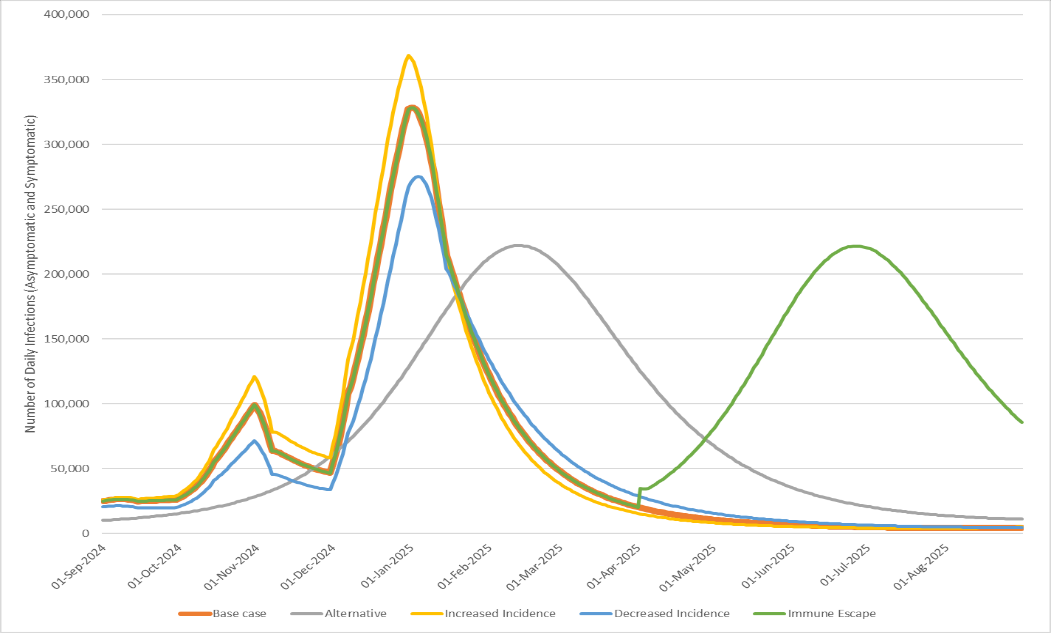

**B: With the Moderna Autumn 2024 Campaign**

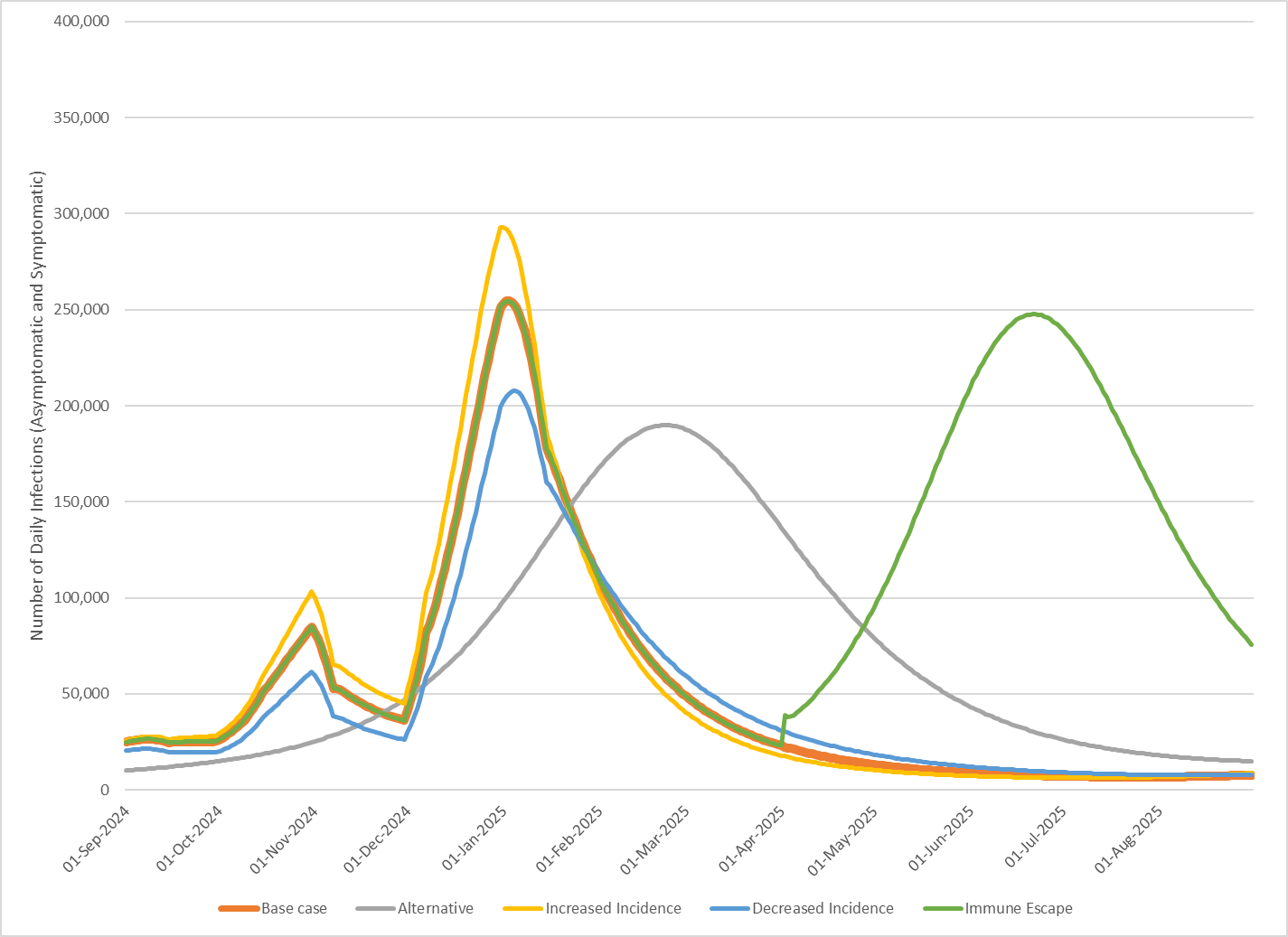

Table 15. Predicted number of hospitalizations with and without the Autumn 2024 vaccine campaign for all incidence scenarios.

| **Incidence Scenario** | **No Autumn 2024 Vaccine** | **Moderna Autumn 2024 Vaccine** |
| --- | --- | --- |
| Base case | 69,254 | 42,703 |
| Alternate | 87,766 | 58,189 |
| Increased transmission | 75,322 | 46,961 |
| Decreased transmission | 61,460 | 37,493 |
| Immune escape | 152,540 | 115,527 |

### 4.3. Scenario analysis: target population age 50+ years. clinical outcomes

Table 16. Scenario analysis: target population age 50+ years,* clinical outcomes with and without the Moderna Autumn 2024 Campaign

| **Outcome** | **No Autumn 2024 Vaccine** | **Moderna Autumn 2024 Vaccine** | **Difference** | **Percent change** |
| --- | --- | --- | --- | --- |
| Number of vaccinations | 0 | 17,091,180 | - | - |
| Symptomatic infections | 8,257,540 | 6,295,885 | -1,961,655 | -24% |
| Hospitalizations | 69,254 | 39,751 | -29,503 | -43% |
| COVID-related deaths | 13,454 | 7,141 | -6,313 | -47% |
| Any long COVID | 306,314 | 233,662 | -72,652 | -24% |
| Severe long COVID | 57,393 | 43,790 | -13,603 | -24% |

*Strategy includes individuals aged 50+ years and those at high-risk under 50 years

### 4.4. Scenario analysis: comparison versus Pfizer-BioNTech Autumn 2024 Campaign, clinical outcomes

Table 17. Scenario analysis: comparison versus the Pfizer-BioNTech Autumn 2024 Campaign, clinical outcomes

| **Outcome** | **Moderna Autumn 2024 Vaccine** | **Pfizer-BioNTech Autumn 2024 Campaign**  **(Kavikondala et al.[50])** | **Difference** | **% change** |
| --- | --- | --- | --- | --- |
| Symptomatic infections | 6,674,735 | 7,319,900 | 645,165 | 10% |
| Hospitalizations | 42,703 | 50,623 | 7,920 | 19% |
| COVID-related deaths | 7,692 | 9,387 | 1,696 | 22% |
| Any long COVID | 247,718 | 271,626 | 23,908 | 10% |
| Severe long COVID | 46,424 | 50,901 | 4,478 | 10% |

Table 18. Scenario analysis: comparison versus the Pfizer-BioNTech Autumn 2024 Campaign, clinical outcomes when coverage of the Pfizer-BioNTech vaccine is increased so that healthcare costs are similar.

| **Clinical Outcomes (Population Total)** | **Comirnaty  (2023/24 VCR) +15%** | **Spikevax  (2023/24 VCR)** | **Difference:  Spikevax – Comirnaty** |
| --- | --- | --- | --- |
| Number of Vaccinations | 21,669,293 | 12,796,178 | -8,873,115 |
| Symptomatic Infections | 6,663,810 | 6,674,735 | 10,925 |
| Hospitalizations | 42,229 | 42,703 | 474 |
| COVID-Related Deaths | 7,739 | 7,692 | -48 |
| Infection-Related Myocarditis | 8,205 | 8,181 | -24 |
| Adverse Event Myocarditis | 62 | 11 | -50 |
| Any Long COVID | 247,310 | 247,718 | 408 |
| Severe Long COVID | 46,348 | 46,424 | 76 |
| Health care treatment costs (millions) | £699 | £700 | £1 |

5. Equations associated with the SEIR Model

### 5.1. Differential equations

The following equations describe the movement of people in the population through the SEIR compartments and the vaccination strata.

**Unvaccinated cohort**

$$S_{t+1,j}^{X}=S_{t,j}^{X}-\lambda_{t,j}^{X}S_{t,j}^{X}-\left( p_{t,j}^{X,S} \right)\mu_{t,j}+\omega_{t,j}^{X}R_{t,j}^{X}-\epsilon_{t,j}$$

$$E_{t+1,j}^{X}=E_{t,j}^{X}+\lambda_{t,j}^{X}S_{t,j}^{X}-\frac{1}{\tau_{E}}E_{t,j}^{X}$$

$$I_{t+1,j}^{X}=I_{t,j}^{X}+\frac{1}{\tau_{E}}E_{t,j}^{X}-\frac{1}{\tau_{I}}I_{t,j}^{X}+\epsilon_{t,j}$$

$$R_{t+1,j}^{X}=R_{t,j}^{X}-\left( p_{t,j}^{X,R} \right)\mu_{t,j}+\frac{1}{\tau_{I}}I_{t,j}^{X}-\omega_{t,j}^{X}R_{t,j}^{X}$$

**Vaccinated cohort**

$$S_{t+1,j}^{V}=S_{t,j}^{V}-\lambda_{t,j}^{V}S_{t,j}^{V}+{\left( p_{t,j}^{X,S} \right)\mu}_{t,j}-\left( p_{t,j}^{V,S} \right)\nu_{t,j}-\left( p_{t,j}^{V,S} \right)p_{t,j}^{V,B2}\nu_{t,j}^{2}-\left( p_{t,j}^{V,S} \right)p_{t,j}^{V,B3}\nu_{t,j}^{3}-\left( p_{t,j}^{V,S} \right)p_{t,j}^{V,B4}\nu_{t,j}^{4}-\left( p_{t,j}^{V,S} \right)p_{t,j}^{V,B5}\nu_{t,j}^{5}-\left( p_{t,j}^{V,S} \right)p_{t,j}^{V,B6}\nu_{t,j}^{6}-\left( p_{t,j}^{V,S} \right)p_{t,j}^{V,B7}\nu_{t,j}^{7}+\omega_{t,j}^{V}R_{t,j}^{V}$$

$$E_{t+1,j}^{V}=E_{t,j}^{V}+\lambda_{t,j}^{V}S_{t,j}^{V}-\frac{1}{\tau_{E}}E_{t,j}^{V}$$

$$I_{t+1,j}^{V}=I_{t,j}^{V}+\frac{1}{\tau_{E}}E_{t,j}^{V}-\frac{1}{\tau_{I}}I_{t,j}^{V}$$

$$R_{t+1,j}^{V}=R_{t,j}^{V}+{\left( p_{t,j}^{X,R} \right)\mu}_{t,j}-\left( p_{t,j}^{V,R} \right)\nu_{t,j}-\left( p_{t,j}^{V,R} \right)p_{t,j}^{V,B2}\nu_{t,j}^{2}-\left( p_{t,j}^{V,R} \right)p_{t,j}^{V,B3}\nu_{t,j}^{3}-\left( p_{t,j}^{V,R} \right)p_{t,j}^{V,B4}\nu_{t,j}^{4}-\left( p_{t,j}^{V,R} \right)p_{t,j}^{V,B5}\nu_{t,j}^{5}-\left( p_{t,j}^{V,R} \right)p_{t,j}^{V,B6}\nu_{t,j}^{6}-\left( p_{t,j}^{V,R} \right)p_{t,j}^{V,B7}\nu_{t,j}^{7}+\frac{1}{\tau_{I}}I_{t,j}^{V}-\omega_{t,j}^{V}R_{t,j}^{V}$$

**First booster cohort**

$$S_{t+1,j}^{B}=S_{t,j}^{B}-\lambda_{t,j}^{B}S_{t,j}^{B}+\left( p_{t,j}^{V,S} \right)\nu_{t,j}-{\left( p_{t,j}^{B,S} \right)\nu}_{t,j}^{2}-{\left( p_{t,j}^{B,S} \right)\left( p_{t,j}^{B,B3} \right)\nu}_{t,j}^{3}-{\left( p_{t,j}^{B,S} \right)\left( p_{t,j}^{B,B4} \right)\nu}_{t,j}^{4}-{\left( p_{t,j}^{B,S} \right)\left( p_{t,j}^{B,B5} \right)\nu}_{t,j}^{5}-{\left( p_{t,j}^{B,S} \right)\left( p_{t,j}^{B,B6} \right)\nu}_{t,j}^{6}-{\left( p_{t,j}^{B,S} \right)\left( p_{t,j}^{B,B7} \right)\nu}_{t,j}^{7}+\omega_{t,j}^{B}R_{t,j}^{B}$$

$$E_{t+1,j}^{B}=E_{t,j}^{B}+\lambda_{t,j}^{B}S_{t,j}^{B}-\frac{1}{\tau_{E}}E_{t,j}^{B}$$

$$I_{t+1,j}^{B}=I_{t,j}^{B}+\frac{1}{\tau_{E}}E_{t,j}^{B}-\frac{1}{\tau_{I}}I_{t,j}^{B}$$

$$R_{t+1,j}^{B}=R_{t,j}^{B}+\left( p_{t,j}^{V,R} \right)\nu_{t,j}-{\left( p_{t,j}^{B,R} \right)\nu}_{t,j}^{2}-{\left( p_{t,j}^{B,R} \right)\left( p_{t,j}^{B,B3} \right)\nu}_{t,j}^{3}-{\left( p_{t,j}^{B,R} \right)\left( p_{t,j}^{B,B4} \right)\nu}_{t,j}^{4}-{\left( p_{t,j}^{B,R} \right)\left( p_{t,j}^{B,B5} \right)\nu}_{t,j}^{5}-{\left( p_{t,j}^{B,R} \right)\left( p_{t,j}^{B,B6} \right)\nu}_{t,j}^{6}-{\left( p_{t,j}^{B,R} \right)\left( p_{t,j}^{B,B7} \right)\nu}_{t,j}^{7}+\frac{1}{\tau_{I}}I_{t,j}^{B}-\omega_{t,j}^{B}R_{t,j}^{B}$$

**Second booster cohort (Spring 2022)**

$$S_{t+1,j}^{B2}=S_{t,j}^{B2}-\lambda_{t,j}^{B2}S_{t,j}^{B2}+\left( p_{t,j}^{V,S} \right)p_{t,j}^{V,B2}\nu_{t,j}^{2}+{\left( p_{t,j}^{B,S} \right)\nu}_{t,j}^{2}-\left( p_{t,j}^{B2,S} \right)\left( p_{t,j}^{B2,B3} \right)\nu_{t,j}^{3}-\left( p_{t,j}^{B2,S} \right)\left( p_{t,j}^{B2,B4} \right)\nu_{t,j}^{4}-\left( p_{t,j}^{B2,S} \right)\left( p_{t,j}^{B2,B5} \right)\nu_{t,j}^{5}-\left( p_{t,j}^{B2,S} \right)\left( p_{t,j}^{B2,B6} \right)\nu_{t,j}^{6}-\left( p_{t,j}^{B2,S} \right)\left( p_{t,j}^{B2,B7} \right)\nu_{t,j}^{7}+\omega_{t,j}^{B2}R_{t,j}^{B2}$$

$$E_{t+1,j}^{B2}=E_{t,j}^{B2}+\lambda_{t,j}^{B2}S_{t,j}^{B2}-\frac{1}{\tau_{E}}E_{t,j}^{B2}$$

$$I_{t+1,j}^{B2}=I_{t,j}^{B2}+\frac{1}{\tau_{E}}E_{t,j}^{B2}-\frac{1}{\tau_{I}}I_{t,j}^{B2}$$

$$R_{t+1,j}^{B2}=R_{t,j}^{B2}+\left( p_{t,j}^{V,R} \right)p_{t,j}^{V,B2}\nu_{t,j}^{2}+{\left( p_{t,j}^{B,R} \right)\nu}_{t,j}^{2}-\left( p_{t,j}^{B2,R} \right)\left( p_{t,j}^{B2,B3} \right)\nu_{t,j}^{3}-\left( p_{t,j}^{B2,R} \right)\left( p_{t,j}^{B2,B4} \right)\nu_{t,j}^{4}-\left( p_{t,j}^{B2,R} \right)\left( p_{t,j}^{B2,B5} \right)\nu_{t,j}^{5}-\left( p_{t,j}^{B2,R} \right)\left( p_{t,j}^{B2,B6} \right)\nu_{t,j}^{6}-\left( p_{t,j}^{B2,R} \right)\left( p_{t,j}^{B2,B7} \right)\nu_{t,j}^{7}+\frac{1}{\tau_{I}}I_{t,j}^{B2}-\omega_{t,j}^{B2}R_{t,j}^{B2}$$

**Third booster cohort (Autumn 2022)**

$$S_{t+1,j}^{B3}=S_{t,j}^{B3}-\lambda_{t,j}^{B3}S_{t,j}^{B3}+\left( \left( p_{t,j}^{V,S} \right)p_{t,j}^{V,B3}+\left( p_{t,j}^{B,S} \right)p_{t,j}^{B,B3}+{\left( p_{t,j}^{B2,S} \right)p}_{t,j}^{B2,B3} \right)\nu_{t,j}^{3}-\left( p_{t,j}^{B3,S} \right)\left( p_{t,j}^{B3,B4} \right)\nu_{t,j}^{4}-\left( p_{t,j}^{B3,S} \right)\left( p_{t,j}^{B3,B5} \right)\nu_{t,j}^{5}-\left( p_{t,j}^{B3,S} \right)\left( p_{t,j}^{B3,B6} \right)\nu_{t,j}^{6}-\left( p_{t,j}^{B3,S} \right)\left( p_{t,j}^{B3,B7} \right)\nu_{t,j}^{7}+\omega_{t,j}^{B3}R_{t,j}^{B3}$$

$$E_{t+1,j}^{B3}=E_{t,j}^{B3}+\lambda_{t,j}^{B3}S_{t,j}^{B3}-\frac{1}{\tau_{E}}E_{t,j}^{B3}$$

$$I_{t+1,j}^{B3}=I_{t,j}^{B3}+\frac{1}{\tau_{E}}E_{t,j}^{B3}-\frac{1}{\tau_{I}}I_{t,j}^{B3}$$

$$R_{t+1,j}^{B3}=R_{t,j}^{B3}+\left( \left( p_{t,j}^{V,R} \right)p_{t,j}^{V,B3}+\left( p_{t,j}^{B,R} \right)p_{t,j}^{B,B3}+{\left( p_{t,j}^{B2,R} \right)p}_{t,j}^{B2,B3} \right)\nu_{t,j}^{3}-\left( p_{t,j}^{B3,R} \right)\left( p_{t,j}^{B3,B4} \right)\nu_{t,j}^{4}-\left( p_{t,j}^{B3,R} \right)\left( p_{t,j}^{B3,B5} \right)\nu_{t,j}^{5}-\left( p_{t,j}^{B3,R} \right)\left( p_{t,j}^{B3,B6} \right)\nu_{t,j}^{6}-\left( p_{t,j}^{B3,R} \right)\left( p_{t,j}^{B3,B7} \right)\nu_{t,j}^{7}+\frac{1}{\tau_{I}}I_{t,j}^{B3}-\omega_{t,j}^{B3}R_{t,j}^{B3}$$

**Fourth booster cohort (Spring 2023)**

$$S_{t+1,j}^{B4}=S_{t,j}^{B4}-\lambda_{t,j}^{B4}S_{t,j}^{B4}+\left( \left( p_{t,j}^{V,S} \right)p_{t,j}^{V,B4}+\left( p_{t,j}^{B,S} \right)p_{t,j}^{B,B4}+{\left( p_{t,j}^{B2,S} \right)p}_{t,j}^{B2,B4}+{\left( p_{t,j}^{B3,S} \right)p}_{t,j}^{B3,B4} \right)\nu_{t,j}^{4}-\left( p_{t,j}^{B4,S} \right)\left( p_{t,j}^{B4,B5} \right)\nu_{t,j}^{5}-\left( p_{t,j}^{B4,S} \right)\left( p_{t,j}^{B4,B6} \right)\nu_{t,j}^{6}-\left( p_{t,j}^{B4,S} \right)\left( p_{t,j}^{B4,B7} \right)\nu_{t,j}^{7}+\omega_{t,j}^{B4}R_{t,j}^{B4}$$

$$E_{t+1,j}^{B4}=E_{t,j}^{B4}+\lambda_{t,j}^{B4}S_{t,j}^{B4}-\frac{1}{\tau_{E}}E_{t,j}^{B4}$$

$$I_{t+1,j}^{B4}=I_{t,j}^{B4}+\frac{1}{\tau_{E}}E_{t,j}^{B4}-\frac{1}{\tau_{I}}I_{t,j}^{B4}$$

$$R_{t+1,j}^{B4}=R_{t,j}^{B4}+\left( \left( p_{t,j}^{V,R} \right)p_{t,j}^{V,B4}+\left( p_{t,j}^{B,R} \right)p_{t,j}^{B,B4}+{\left( p_{t,j}^{B2,R} \right)p}_{t,j}^{B2,B4}+{\left( p_{t,j}^{B3,R} \right)p}_{t,j}^{B3,B4} \right)\nu_{t,j}^{4}-\left( p_{t,j}^{B4,R} \right)\left( p_{t,j}^{B4,B5} \right)\nu_{t,j}^{5}-\left( p_{t,j}^{B4,R} \right)\left( p_{t,j}^{B4,B6} \right)\nu_{t,j}^{6}-\left( p_{t,j}^{B4,R} \right)\left( p_{t,j}^{B4,B7} \right)\nu_{t,j}^{7}+\frac{1}{\tau_{I}}I_{t,j}^{B4}-\omega_{t,j}^{B4}R_{t,j}^{B4}$$

**Fifth booster cohort (Autumn 2023)**

$$S_{t+1,j}^{B5}=S_{t,j}^{B5}-\lambda_{t,j}^{B5}S_{t,j}^{B5}+\left( \left( p_{t,j}^{V,S} \right)p_{t,j}^{V,B5}+\left( p_{t,j}^{B,S} \right)p_{t,j}^{B,B5}+{\left( p_{t,j}^{B2,S} \right)p}_{t,j}^{B2,B5}+{\left( p_{t,j}^{B3,S} \right)p}_{t,j}^{B3,B5}+{\left( p_{t,j}^{B4,S} \right)p}_{t,j}^{B4,B5} \right)\nu_{t,j}^{5}-\left( p_{t,j}^{B5,S} \right)\left( p_{t,j}^{B5,B6} \right)\nu_{t,j}^{6}-\left( p_{t,j}^{B5,S} \right)\left( p_{t,j}^{B5,B7} \right)\nu_{t,j}^{7}+\omega_{t,j}^{B5}R_{t,j}^{B5}$$

$$E_{t+1,j}^{B5}=E_{t,j}^{B5}+\lambda_{t,j}^{B5}S_{t,j}^{B5}-\frac{1}{\tau_{E}}E_{t,j}^{B5}$$

$$I_{t+1,j}^{B5}=I_{t,j}^{B5}+\frac{1}{\tau_{E}}E_{t,j}^{B5}-\frac{1}{\tau_{I}}I_{t,j}^{B5}$$

$$R_{t+1,j}^{B5}=R_{t,j}^{B5}+\left( \left( p_{t,j}^{V,R} \right)p_{t,j}^{V,B5}+\left( p_{t,j}^{B,R} \right)p_{t,j}^{B,B5}+{\left( p_{t,j}^{B2,R} \right)p}_{t,j}^{B2,B5}+{\left( p_{t,j}^{B3,R} \right)p}_{t,j}^{B3,B5}+{\left( p_{t,j}^{B4,R} \right)p}_{t,j}^{B4,B5} \right)\nu_{t,j}^{5}-\left( p_{t,j}^{B5,R} \right)\left( p_{t,j}^{B5,B6} \right)\nu_{t,j}^{6}-\left( p_{t,j}^{B5,R} \right)\left( p_{t,j}^{B5,B7} \right)\nu_{t,j}^{7}+\frac{1}{\tau_{I}}I_{t,j}^{B5}-\omega_{t,j}^{B5}R_{t,j}^{B5}$$

**Sixth booster cohort (Spring 2024)**

$$S_{t+1,j}^{B6}=S_{t,j}^{B6}-\lambda_{t,j}^{B6}S_{t,j}^{B6}+\left( \left( p_{t,j}^{V,S} \right)p_{t,j}^{V,B6}+\left( p_{t,j}^{B,S} \right)p_{t,j}^{B,B6}+{\left( p_{t,j}^{B2,S} \right)p}_{t,j}^{B2,B6}+{\left( p_{t,j}^{B3,S} \right)p}_{t,j}^{B3,B6}+{\left( p_{t,j}^{B4,S} \right)p}_{t,j}^{B4,B6}+{\left( p_{t,j}^{B5,S} \right)p}_{t,j}^{B5,B6} \right)\nu_{t,j}^{6}-\left( p_{t,j}^{B6,S} \right)\left( p_{t,j}^{B6,B7} \right)\nu_{t,j}^{7}+\omega_{t,j}^{B6}R_{t,j}^{B6}$$

$$E_{t+1,j}^{B6}=E_{t,j}^{B6}+\lambda_{t,j}^{B6}S_{t,j}^{B6}-\frac{1}{\tau_{E}}E_{t,j}^{B6}$$

$$I_{t+1,j}^{B6}=I_{t,j}^{B6}+\frac{1}{\tau_{E}}E_{t,j}^{B6}-\frac{1}{\tau_{I}}I_{t,j}^{B6}$$

$$R_{t+1,j}^{B6}=R_{t,j}^{B6}+\left( \left( p_{t,j}^{V,R} \right)p_{t,j}^{V,B6}+\left( p_{t,j}^{B,R} \right)p_{t,j}^{B,B6}+{\left( p_{t,j}^{B2,R} \right)p}_{t,j}^{B2,B6}+{\left( p_{t,j}^{B3,R} \right)p}_{t,j}^{B3,B6}+{\left( p_{t,j}^{B4,R} \right)p}_{t,j}^{B4,B6}+{\left( p_{t,j}^{B5,R} \right)p}_{t,j}^{B5,B6} \right)\nu_{t,j}^{6}-\left( p_{t,j}^{B6,R} \right)\left( p_{t,j}^{B6,B7} \right)\nu_{t,j}^{7}+\frac{1}{\tau_{I}}I_{t,j}^{B6}-\omega_{t,j}^{B6}R_{t,j}^{B6}$$

**Eighth booster cohort (Autumn 2024)**

$$S_{t+1,j}^{B7}=S_{t,j}^{B7}-\lambda_{t,j}^{B7}S_{t,j}^{B7}+\left( \left( p_{t,j}^{V,S} \right)p_{t,j}^{V,B7}+\left( p_{t,j}^{B,S} \right)p_{t,j}^{B,B7}+{\left( p_{t,j}^{B2,S} \right)p}_{t,j}^{B2,B7}+{\left( p_{t,j}^{B3,S} \right)p}_{t,j}^{B3,B7}+{\left( p_{t,j}^{B4,S} \right)p}_{t,j}^{B4,B7}+{\left( p_{t,j}^{B5,S} \right)p}_{t,j}^{B5,B7}+{\left( p_{t,j}^{B6,S} \right)p}_{t,j}^{B6,B7} \right)\nu_{t,j}^{7}+\omega_{t,j}^{B7}R_{t,j}^{B7}$$

$$E_{t+1,j}^{B7}=E_{t,j}^{B7}+\lambda_{t,j}^{B7}S_{t,j}^{B7}-\frac{1}{\tau_{E}}E_{t,j}^{B7}$$

$$I_{t+1,j}^{B7}=I_{t,j}^{B7}+\frac{1}{\tau_{E}}E_{t,j}^{B7}-\frac{1}{\tau_{I}}I_{t,j}^{B7}$$

$$R_{t+1,j}^{B7}=R_{t,j}^{B7}+\left( \left( p_{t,j}^{V,R} \right)p_{t,j}^{V,B7}+\left( p_{t,j}^{B,R} \right)p_{t,j}^{B,B7}+{\left( p_{t,j}^{B2,R} \right)p}_{t,j}^{B2,B7}+{\left( p_{t,j}^{B3,R} \right)p}_{t,j}^{B3,B7}+{\left( p_{t,j}^{B4,R} \right)p}_{t,j}^{B4,B7}+{\left( p_{t,j}^{B5,R} \right)p}_{t,j}^{B5,B7}+{\left( p_{t,j}^{B6,R} \right)p}_{t,j}^{B6,B7} \right)\nu_{t,j}^{7}+\frac{1}{\tau_{I}}I_{t,j}^{B7}-\omega_{t,j}^{B7}R_{t,j}^{B7}$$

**For t = 1,**

$S_{1,j}^{X}$ = Initial number of susceptible individuals

$I_{1,j}^{X}$= Initial number of infectious individuals = $\epsilon_{1,j}$

$$E_{1,j}^{X}=R_{1,j}^{X}=0$$

$$S_{1,j}^{V}=E_{1,j}^{V}=I_{1,j}^{V}=R_{1,j}^{V}=0$$

$$S_{1,j}^{B}=E_{1,j}^{B}=I_{1,j}^{B}=R_{1,j}^{B}=0$$

$$S_{1,j}^{B2}=E_{1,j}^{B2}=I_{1,j}^{B2}=R_{1,j}^{B2}=0$$

$$S_{1,j}^{B3}=E_{1,j}^{B3}=I_{1,j}^{B3}=R_{1,j}^{B3}=0$$

$$S_{1,j}^{B4}=E_{1,j}^{B4}=I_{1,j}^{B4}=R_{1,j}^{B4}=0$$

$$S_{1,j}^{B5}=E_{1,j}^{B5}=I_{1,j}^{B5}=R_{1,j}^{B5}=0$$

$$S_{1,j}^{B6}=E_{1,j}^{B6}=I_{1,j}^{B6}=R_{1,j}^{B6}=0$$

$$S_{1,j}^{B7}=E_{1,j}^{B7}=I_{1,j}^{B7}=R_{1,j}^{B7}=0$$

Notes, superscripts:

*X, V, B, B2, B3, B4, B5, B6, and B7 represent the unvaccinated, vaccinated, first booster, second booster (Spring 2022), third booster (Autumn 2022), fourth booster (Spring 2023), fifth booster (Autumn 2023), sixth booster (Spring 2024), and seventh booster (Autumn 2024) cohorts, respectively.*

Notes, subscripts:

*t* = time (i.e., day of analysis)

*j* = age group (number 1 to 9)

**Definitions**

$S_{j}^{X}, S_{j}^{V}, S_{j}^{B}, S_{j}^{B2},S_{j}^{B3},S_{j}^{B4},S_{j}^{B5},S_{j}^{B6},S_{j}^{B7}$ represent the proportion of susceptibles in age group *j* in cohort *X*, *V, B,* *B2, B3*, B4, B5, B6, or *B7*

The compartments with superscript *X*, *V*, *B, B1, B2, B3, B4, B5, B6, and B7* represent the unvaccinated, vaccinated, and boosted cohorts, respectively.

$E_{j}^{Z}$ represent exposed, but not yet infectious, individuals in age group *j* in cohort *Z* (*X, V, B, B2, B3, B4, B5, B6, or B7*).

$I_{j}^{Z}$ represent infectious individuals in age group *j* in cohort *Z* (*X, V, B, B2, B3, B4, B5, B6, or B7*).

$R_{j}^{Z}$ represent immune individuals in age group *j* in cohort *Z* (*X, V, B, B2, B3, B4, B5, B6, or B7*).

$\lambda_{t,j}^{*}$is the age-group specific force of infection (see section below)

$\frac{1}{\tau_{E}}$ is the rate of loss of latency

$\frac{1}{\tau_{I}}$ is the rate of loss of infectiousness

$\mu_{t,j}$ is the proportion receiving a (primary series) vaccination on day *t* in age group *j*

$\nu_{t,j}$ is the proportion receiving a first booster on day *t* in age group *j*

$\nu_{t,j}^{2}$ is the proportion receiving a second booster (Spring 2022) on day *t* in age group *j*

$\nu_{t,j}^{3}$ is the proportion receiving a third booster (Autumn 2022) on day *t* in age group *j*

$\nu_{t,j}^{4}$ is the proportion receiving a fourth booster (Spring 2023) on day *t* in age group *j*

$\nu_{t,j}^{5}$ is the proportion receiving a fifth booster (Autumn 2023) on day *t* in age group *j*

$\nu_{t,j}^{6}$ is the proportion receiving a sixth booster (Spring 2024) on day *t* in age group *j*

$\nu_{t,j}^{7}$ is the proportion receiving a seventh booster (Autumn 2024) on day *t* in age group *j*

$p_{t,j}^{Z,Z^{*}}$ is the proportion receiving a booster in cohort *Z** from cohort *Z* on day *t* in age group *j*

*{e.g.,* $p_{t,j}^{V,B5}$ is the proportion receiving a fifth booster from cohort *V* on day *t* in age group *j}*

Note: $p_{t,j}^{V,B2}+p_{t,j}^{B,B2}=1$

Note: $p_{t,j}^{V,B3}+p_{t,j}^{B,B3}+p_{t,j}^{B2,B3}=1$

Note: $p_{t,j}^{V,B4}+p_{t,j}^{B,B4}+p_{t,j}^{B2,B4}+p_{t,j}^{B3,B4}=1$

Note: $p_{t,j}^{V,B5}+p_{t,j}^{B,B5}+p_{t,j}^{B2,B5}+p_{t,j}^{B3,B5}+p_{t,j}^{B4,B5}=1$

Note: $p_{t,j}^{V,B6}+p_{t,j}^{B,B6}+p_{t,j}^{B2,B6}+p_{t,j}^{B3,B6}+p_{t,j}^{B4,B6}+p_{t,j}^{B5,B6}=1$

Note: $p_{t,j}^{V,B7}+p_{t,j}^{B,B7}+p_{t,j}^{B2,B7}+p_{t,j}^{B3,B7}+p_{t,j}^{B4,B7}+p_{t,j}^{B5,B7}+p_{t,j}^{B6,B7}=1$

$p_{t,j}^{Z,S}$ is the proportion of the booster cohort *Z in the S* compartment out of the total proportion of the booster cohort *Z in the S and R* compartments on day *t* in age group *j*

$p_{t,j}^{Z,R}$ is the proportion of the booster cohort *Z in the R* compartment out of the total proportion of the booster cohort *Z in the S and R* compartments on day *t* in age group *j*

Note: $p_{t,j}^{Z,S}+p_{t,j}^{Z,R}=1$

$\omega_{t,j}^{Z}$ is the natural immunity waning rate on day *t* in age group *j* in cohort Z (*X, V, B, B2, B3, B4, B5, B6, or B7)*

$\epsilon_{t,j}$ is the proportion of external cases on day *t* in age group *j*

### 5.2. Force of infection (unvaccinated)

$$\lambda_{t,i}^{*}=\left( {Overall Scaling Factor}_{t} \right)\times\left[ \beta_{t}\sum_{j=1}^{9} \sum_{Z=\left\{ X,V,B,\ldots,B7 \right\}} c_{ij}I_{t,j}^{Z} \right]$$

$\lambda_{t,i}^{*}$is the age-group specific force of infection at time *t* for age group *i*

${Overall Scaling Factor}_{t}$at time *t* is defined in section 1.2.2.

$\beta_{t}$ is the transmissibility parameter at time *t*

$c_{ij}$ is the rate at which individuals in age group *i* make contact with those in age group *j*

$I_{t,j}^{Z}$ represents the infectious individuals in cohort *Z* (*X, V, B, B2, B3, B4, B5, B6, or B7*).at time *t* for age group *j*.

### 5.3. Force of infection (vaccinated)

For the vaccinated cohorts, the force of infection calculation is adjusted based on the VE in the cohort:

$$\lambda_{t,i}^{Z}=\left( 1-{VE}_{t,i}^{X} \right)\times\left( {Overall Scaling Factor}_{t} \right)\times\left[ \beta_{t}\sum_{j=1}^{9} \sum_{Z=\left\{ X,V,B,\ldots,B7 \right\}} c_{ij}I_{t,j}^{Z} \right]$$

$\lambda_{t,i}^{Z}$is the age-group specific force of infection at time *t* for age group *i* in cohort *Z* (*X, V, B, B2, B3, B4, B5, B6, or B7*).

${VE}_{t,i}^{Z}$ is the vaccine effectiveness at time *t* for age group *i* in cohort *Z* (*X, V, B, B2, B3, B4, B5, B6, or B7*). The vaccine effectiveness at time *t* is defined in the next section.

### 5.4. Daily vaccine effectiveness calculations

If no one is vaccinated in the cohort and age group, a VE value of zero is assumed (i.e., when day *t* is less than the day of the beginning of the vaccination period). Once people have been vaccinated in the vaccination cohort and age group, the average vaccine effectiveness on day *t* is calculated as:

$${VE}_{t,i}^{Z}=\frac{\left[ \begin{aligned} \left( Number newly vaccinated on day t\times initial VE \right)+ \\ \left( \left( {VE Drop}_{t} \right)\left( Number previously vaccinated on day t\times({VE}_{t-1,i}^{Z}-daily waning rate \right) \right) \end{aligned} \right]}{\left[ Total number in cohort Z on day t \right]}$$

${VE}_{t,i}^{Z}$ is the vaccine effectiveness at time *t* for age group *i* in cohort *Z* (*X, V, B, B2, B3, B4, B5, B6, or B7*).

If the term $\left( {VE}_{t-1,i}^{Z}-daily waning rate \right)$ falls below zero, we assume a value of zero instead.

The term $\left( {VE Drop}_{t} \right)$ represents a drop in the vaccine effectiveness when a new strain with immune escape enters the population. There are only a few days in which this drop occurs over the course of the time horizon: if a new variant with immune escape emerges, the impact is assumed to happen on the individual days. Apart from those days, the value of the term $\left( {VE Drop}_{t} \right)$ is assumed to be one, corresponding to no impact on the average VE calculation.

### 5.5. Calculation of incremental effectiveness against hospitalization

For each vaccination cohort, age group, and day, we define the following vaccine effectiveness variables and relationship between the variables. The superscripts and subscripts for vaccination cohort, age group, and day are removed for clarity.

**Definitions**

${VE}_{1}$ = Vaccine effectiveness against infection

${VE}_{2}$ = ‘Total’ Vaccine effectiveness against hospitalization

${VE}_{2}^{*}$= ‘Additional’ Vaccine effectiveness against hospitalization

We assume ${VE}_{2}^{*}=0$ if there is no additional benefit against hospitalization

**Define**

$$\left[ 1-{VE}_{2} \right]= \left[ 1-{VE}_{1} \right]\times\left[ 1-{VE}_{2}^{*} \right]$$

Isolate and solve for ${VE}_{2}^{*}$

$$\left[ 1-{VE}_{2}^{*} \right]=\frac{\left[ 1-{VE}_{2} \right]}{\left[ 1-{VE}_{1} \right]}$$

$${VE}_{2}^{*}= 1- \frac{\left[ 1-{VE}_{2} \right]}{\left[ 1-{VE}_{1} \right]}$$

**Probabilities in an unvaccinated cohort**

Probability of COVID-19 infection in an unvaccinated cohort

$$p\left( COVID|UnVac \right)$$

Probability that a COVID-19 infection requires hospitalization in an unvaccinated cohort

$$p\left( Hosp|COVID,UnVac \right)$$

Proportion of an unvaccinated cohort with a COVID-19 infection that requires hospitalization

$$p\left( Hosp,COVID|UnVac \right)= p\left( Hosp|COVID,UnVac \right) \times p\left( COVID|UnVac \right)$$

**Probabilities in a vaccinated cohort**

Probability of COVID-19 infection in a vaccinated cohort

$$p\left( COVID|Vac \right)= \left[ 1-{VE}_{1} \right]\times p\left( COVID|UnVac \right)$$

Probability that an COVD-19 infection requires hospitalization in a vaccinated cohort

$$p\left( Hosp|COVID,Vac \right)= \left[ 1-{VE}_{2}^{*} \right]\times p\left( Hosp|COVID,UnVac \right)$$

Proportion of a vaccinated cohort with an COVID infection that requires hospitalization

$$p\left( Hosp,COVID|Vac \right)= p\left( Hosp|COVID,Vac \right) \times p\left( COVID|Vac \right)$$
